# Childhood maltreatment influences adult brain structure through its effects on immune, metabolic and psychosocial factors

**DOI:** 10.1101/2023.06.15.23291420

**Authors:** Sofia C. Orellana, Richard A.I. Bethlehem, Ivan Simpson-Kent, Anne-Laura van Harmelen, Petra E. Vértes, Edward T. Bullmore

## Abstract

Childhood maltreatment (CM) leads to a lifelong susceptibility to mental ill-health which might be reflected by it’s effects on adult brain structure, perhaps indirectly mediated by its effects on adult metabolic, immune, and psychosocial systems. Indexing these systemic factors via body mass index (BMI), C-reactive protein (CRP) and rates of adult trauma (AT), respectively, we tested three hypotheses: (H1) CM has direct or indirect causal effects on adult trauma, BMI and CRP; (H2) adult trauma, BMI and CRP are all independently related to adult brain structure; and (H3) effects of CM on adult brain structure are mediated by its effects on adult trauma, BMI and CRP. Using path analysis and data from N=116,887 participants in UK Biobank we find that CM is related to greater BMI and AT levels, and that only these two variables mediate CM’s effects on CRP [H1]. Regression analyses on the UKB MRI sub-sample (N=21,738) revealed that greater CRP and BMI were both related to a spatially convergent pattern of cortical effects (Spearman’s *ρ*=0.87) characterised by fronto-occipital increases and temporo-parietal reductions in thickness, and that AT is related to lower subcortical volumes [H2]. Finally, path models indicated that CM has indirect effects in a subset of brain regions through its influence on BMI, CRP and AT [H3]. Results provide evidence that childhood maltreatment can influence brain structure decades after exposure by increasing individual risk towards adult trauma, obesity or inflammation.

## Introduction

Childhood maltreatment (CM) is commonly understood to comprise experiences of emotional, physical and sexual abuse, or emotional and physical neglect, in the first 15 years of life [31]. Individuals affected by CM have an increased lifetime risk of mental ill-health [57] representing approximately 30% of psychiatric patients worldwide [49], andcharacterized clinically by an increased recurrence, earlier onset and greater severity of mental health symptoms [55, 57]. This lifelong vulnerability to mental ill-health is likely conferred by structural brain alterations commonly present in individuals exposed to early adversity [71]. Here, we aim to shed light on the mediating pathways that link exposure to childhood maltreatment to alterations in brain structure many decades later.

It is well-established that early adversity has several effects within the highly inter-related domains of immune, metabolic and psychosocial function. In both prospective human studies and animal models, childhood maltreatment precedes the onset of low-grade chronic inflammation in the periphery, even in the absence of infection [21, 41]. Blood inflammatory status has typically been indexed by serum concentration of pro-inflammatory cytokines and acute phase proteins, such as C-reactive protein (CRP), which are molecular markers of activation of the innate immune system [48, 7]. Increased blood cytokine and CRP levels have been shown to predict the onset of psychopathology across several disorders [78, 79], and are characteristically increased in patients with a history of childhood adversity [14, 17].

Metabolically, early life maltreatment is linked to a higher risk of obesity and its associated disorders, such as diabetes and cardiovascular disease [20, 6]. Obesity is a chronic pro-inflammatory factor, [75] as adipose tissue includes many innate immune cells which can release pro-inflammatory cytokines into the circulation [60]. Psychosocially, childhood maltreatment is strongly predictive of re-victimization in the form of adult traumatic experiences involving interpersonal conflict, assault or material precariousness [105, 13]. Physiological responses to repeated social stress include chronic activation of the hypothalamic-pituitary-adrenal (HPA) axis and compensatory down-regulation of anti-inflammatory signalling mediated via the glucocorticoid receptor (GR) [45, 85]. Hypothetically, therefore, long-term metabolic and social effects of CM could contribute to increased inflammation in adult survivors of early adversity.

Alterations in brain structure have also been associated with inflammation, obesity and adult trauma. Global reductions in brain volume and decreased cortical thickness in the insula and pre-frontal cortex have been linked to increased blood CRP levels [37, 74], and increased CRP has also been associated with cerebral atrophy in diverse neurodegenerative disorders [100]. Greater body-mass-index (BMI) is related to decreased global brain volume [39] and visceral fat content has a U-shaped relationship with cortical thickness across all lobes [16], in line with reports of widespread decreases in region-specific cortical thickness associated with higher BMI [103]. Adult trauma exposure has been associated with hippocampal volume reduction [108, 58] as well as changes in thickness of prefrontal and temporo-parietal cortical areas [47, 88]. Consequently, it is possible that early adversity has long term influences on brain structure through its preceding impact on immune, metabolic, nervous and psychosocial systems.

We therefore sought to shed light on the long term consequences of childhood maltreatment on psychophysiological systems and their interactions. We reasoned that there could be a chain of causal connections linking exposure to childhood maltreatment to alterations in adult brain structure via the intermediate effects of childhood maltreatment on risks for obesity, inflammation and adult trauma. On this basis, we tested the following hypotheses: (H1) childhood maltreatment has indirect effects on CRP through its direct effects on BMI and adult trauma; (H2) adult trauma, BMI and CRP are all independently related to adult brain structure; and (H3) effects of CM on adult brain structure are mediated by its effects on adult trauma, BMI and CRP.

## Results

### Sample

We test all hypotheses on the *N* =21,738 participants (UKB MRI sample) who had complete, quality controlled T1-weighted and T2-weighted whole brain MRI data, as well as data on adult trauma, BMI, and CRP available. We additionally replicate H1 on a larger sample (*N* =116,887) of participants who met the relevant inclusion criteria for this study but lacked imaging data (Table 1).

**Table 1:** Demographic and clinical data on UKB sample and sub-sample with MRI data available. Mean and standard deviation (in brackets) are reported for each variable, unless otherwise specified.

|  | UKB sample | 95% CI | UKB MRI sub-sample | 95% CI |
| --- | --- | --- | --- | --- |
| N | 116,887 | 100%, 100% | 21,738 | 100%, 100% |
| Female n(%) | 65,715 (56%) | - | 11,684 (54%) | - |
| Age at baseline | 56 (8) | 56, 56 | 55 (7) | 55, 55 |
| Age at MRI | - | - | 64 (7) | 64, 64 |
| CRP, mg/L 10 | 2.29 (4.02) | 2.3, 2.3 | 2.03 (3.53) | 2.0, 2.1 |
| CRP, log | 0.21 (1.05) | 0.20, 0.22 | 0.11 (1.02) | 0.10, 0.12 |
| Childhood Maltreatment | 1.76 (2.41) | 1.7, 1.8 | 1.70 (2.31) | 1.7, 1.7 |
| Adult Trauma | 2.09 (2.51) | 2.1, 2.1 | 1.93 (2.37) | 1.9, 2.0 |
| Body Mass Index (kg/m<sup>2</sup>) | 26.8 (4.6) | 27, 27 | 26.5 (4.2) | 26, 27 |

### Relationships between childhood maltreatment, adult trauma, BMI and CRP

We used path analysis to examine the magnitude of hypothetically directed relationships between these variables as defined by **H1**: childhood maltreatment has effects on adult inflammation (CRP) mediated directly or indirectly by its direct effects on BMI and AT. The correlation matrix demonstrated small-to-moderate positive correlations between each pair of the four model variables (Fig 1A). The strongest correlations were between CRP and BMI (*r* = 0.43), and between CM and AT (*r* = 0.31). All correlations were statistically significant, even when smaller, reflecting the large sample size.

**Figure 1:**
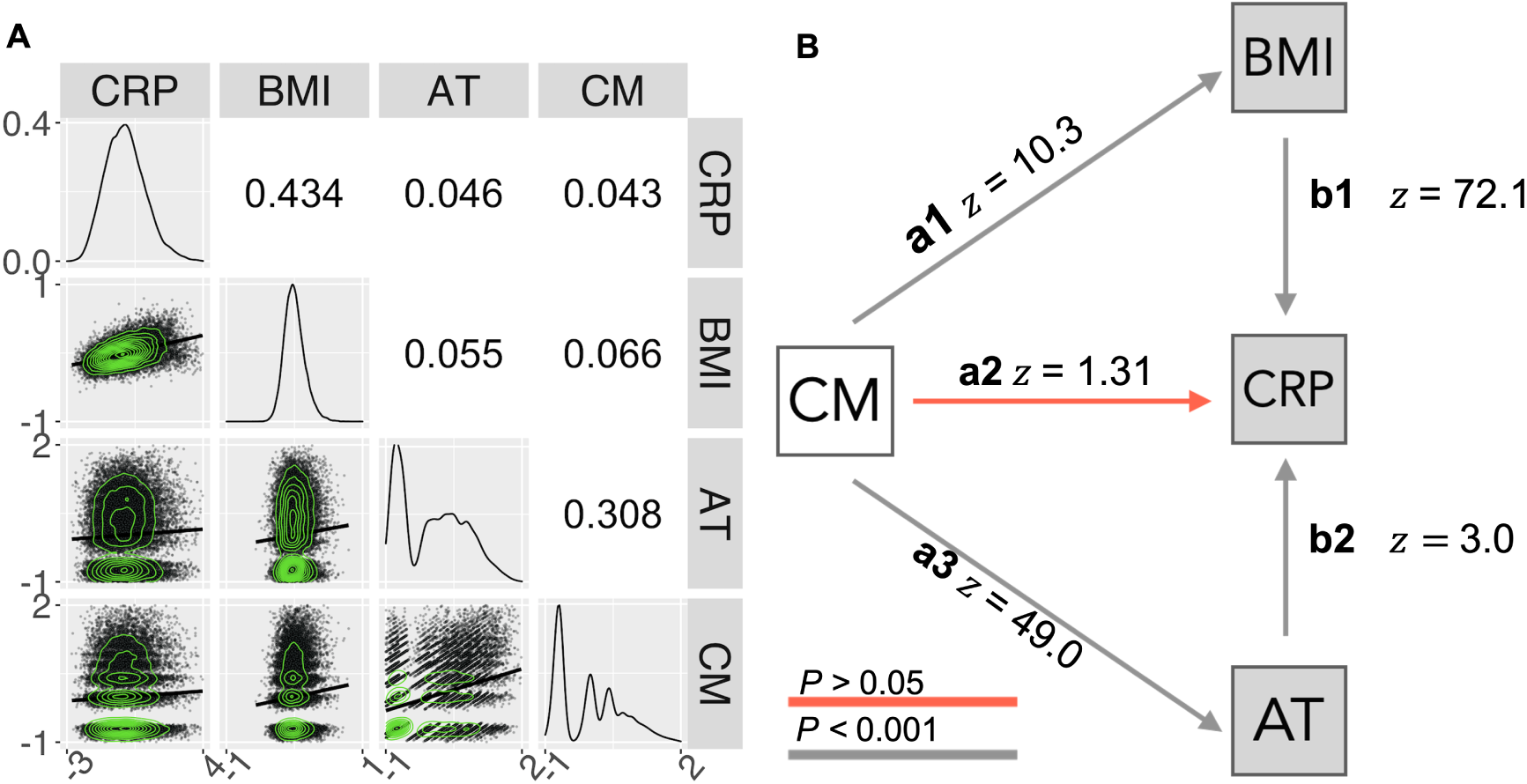
Relationships between childhood maltreatment, adult trauma, BMI and CRP. A. Correlation matrix representing pair-wise Spearman’s correlations (upper triangle) and scatterplots of the relationships between each pair of variables, with solid lines indicating fitted linear regression models (lower triangle). The diagonal represents the probability density function for each variable. All correlations were significantly greater than zero, with FDR *≤* 0.05. **B.** Path diagram representing direct effects of retrospectively ascertained CM (white) on the contemporaneously measured adult (log-transformed) variables, AT, CRP, and BMI (grey). Standardized path coefficients are given as Wald (*z*) statistics.

The model fit to this correlation matrix included direct effects of retrospectively assessed childhood maltreatment on all 3 adult variables (denoted *CM → BMI* (*a1*), *CM → AT* (*a3*), and *CM → CRP* (*a2*); as well as direct effects of AT and BMI on contemporaneously measured CRP (denoted *BMI → CRP (*b1) and *AT → CRP (*b2). The fit for this model was good: *SRMR* = 0.012; *CFI* = 0.996; *RMSEA* = 0.035.

The estimated path coefficients and their standard errors indicated that childhood maltreatment significantly predicted both higher adult BMI (*CM → BMI, z* = 10.343*, β* = 0.072*, P <* 0.001) and greater adult trauma (*CM → AT, z* = 48.882*, β* = 0.315*, P <* 0.001). Note that *z* values repesent Wald *z*’s: *β* coefficients standardized by their standard errors. The direct effects of adult trauma and BMI on CRP were also significant: higher BMI (*BMI → CRP, z* = 72.047*, β* = 0.434*, P <* 0.001) and higher adult trauma (*AT → CRP, z* = 2.999*, β* = 0.019*, P* = 0.003) were both predictive of higher blood concentration of CRP (Fig 1A). When these contemporaneous effects on CRP were taken into account, the path coefficient representing a direct effect of childhood maltreatment on adult CRP was not significant (*CM → CRP, z* = 1.313*, β* = 0.008*, P* = 0.189) (Table ST3). However, indirect effects of childhood maltreatment on CRP, mediated by its direct effects on adult trauma and BMI, were significant: *CM → BMI → CRP, z* = 10.257*, β* = 0, 031*, P <* 0.001 (*a1*b1*); and *CM → AT → CRP, z* = 3.000*, β* = 0.006*, P* = 0.003 (*a3*b2*) respectively. To test if the previously reported relationship between CM and CRP [7] disappeared due to the inclusion of indirect effects, we examined the relationship between CM and CRP with a simple linear regression, which yielded significant results (*β*(*SE*) = 0.03177(0.00470)*, t* = 6.757*, P <* 0.0001).

This analysis of data from the UKB MRI sample was repeated using the larger dataset available on the UKB sample (ST4). In this case, the direct effect of CM on CRP was small but statistically significant: *CM → CRP, β* = 0.0095*, z* = 3.431*, P <* 0.001); see Supplemental Information SI 1.3, Fig. S4B and Table ST4 for details.

### Effects of adult trauma, BMI and CRP on cortical thickness and subcortical volumes

To test **(H2)**, -i.e. adult trauma, BMI and CRP are all independently related to adult brain structure-these three variables were each treated separately as predictors in three different linear regression models with cortical thickness (or subcortical volume) as the dependent variable at each of 180 cortical areas (or 7 subcortical structures) (Fig. 2).

**Figure 2:**
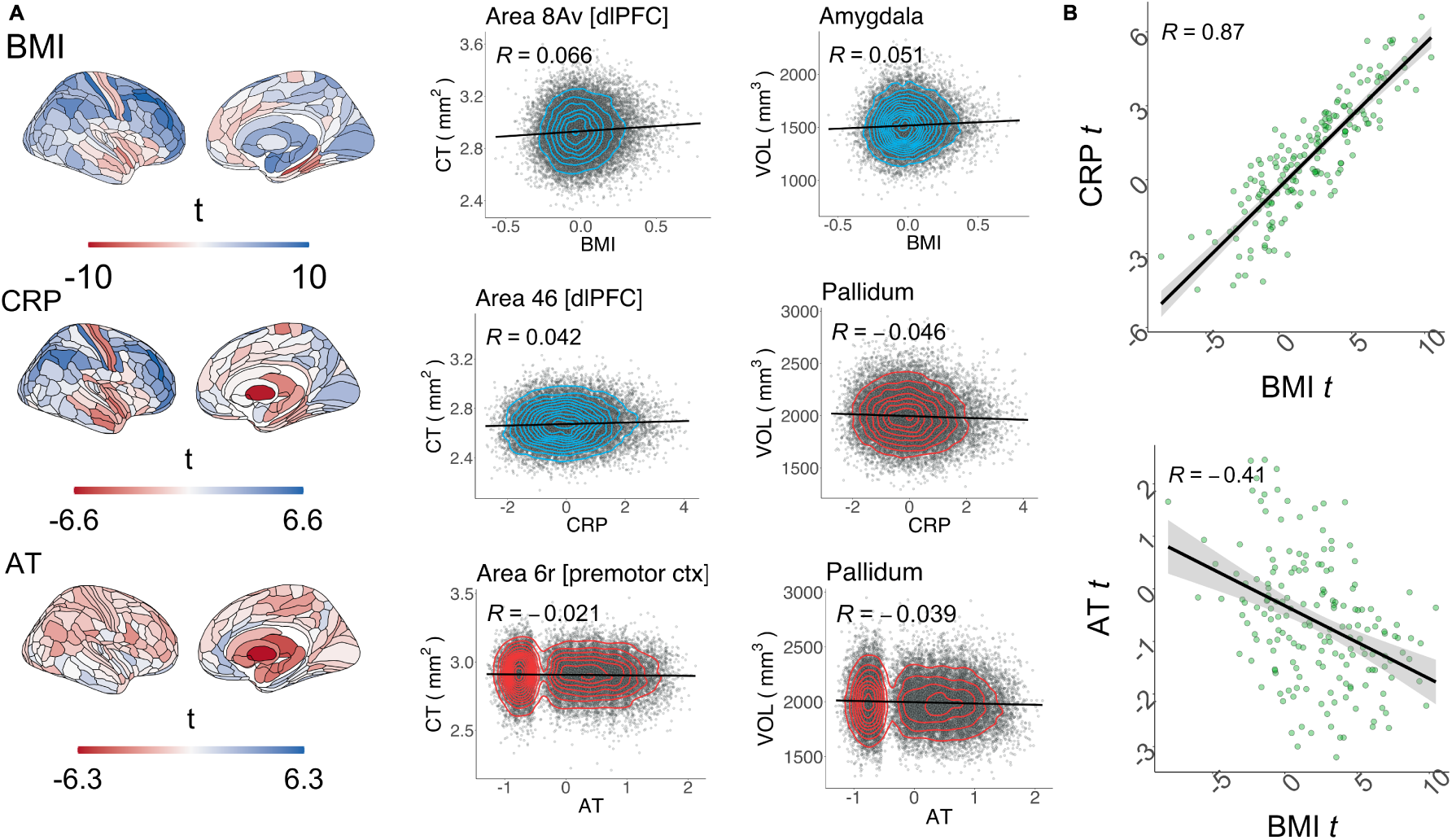
Effects of adult trauma, BMI and CRP on adult cortical thickness and subcortical volumes. A, left column. Brain maps of independent linear relationships between adult trauma (AT), CRP or BMI and cortical thickness (CT) and subcortical volume. Each map shows the anatomical distribution of unthresholded effects (*t* values); for corresponding maps thresholded for FDR-corrected significance see Figure S8; for maps of standardized regression *β* coefficients see S10A & B. **A, right column** Illustrative scatterplots of the relationship between each independent variable and the cortical area or subcortical structure most strongly associated with it. Color indicates the sign of association as either positive (blue) or negative (red). Cortical areas are labelled by their specific areal nomenclature and corresponding regional grouping, as defined by the Glasser template [34]. **B, top panel** Scatterplot of the effect (*t*-value) of BMI (*x*-axis) versus the effect of CRP (*t*-value; *y*-axis) on cortical thickness; each point represents one of 180 cortical areas; Spearman’s correlation *ρ* = 0.87*, P_spin_ <* 0.0001 over all areas; solid line is the regression of *t_BMI_* on *t_CRP_* . **B, bottom panel** Scatter-plot of effect of BMI on CT (*x*-axis) versus effect of AT on CT (*y*-axis); Spearman’s *ρ* = *−*0.41, *P_spin_* = 0.0009

At the cortical level, BMI and CRP were each associated with very similar anatomical patterns of cortical thickness variation. The unthresholded maps of linear regression coefficients of BMI and CRP effects on cortical thickness were highly correlated across all cortical areas and robust to spatial autocorrelation as controlled with a spin test (Spearman’s *ρ* = 0.87, *P_spin_ <* 0.0001); Figure 2B. When the effects of CRP and BMI were separately tested for statistical significance at each brain region, with FDR *≤* 0.05, an extensively overlapping set of regions was identified as significantly associated with both CRP and BMI (Figure S8).

Higher BMI was predictive of significantly increased cortical thickness in 92 areas, with the top 10 largest effects in dorsolateral-prefrontal cortex, primary somatosensory and motor cortices, pre-motor and inferior frontal cortices; as well as significantly decreased cortical thickness in 16 areas, with the strongest decreases located in medial temporal, insular and frontal opercular, primary somatosensory and motor, anterior cingulate and medial prefrontal, association auditory and orbitofrontal cortices (see ST6).

Likewise, higher CRP was predictive of significantly increased cortical thickness in 50 areas, with the top 10 largest significant effects located in dorsolateral-prefrontal, early somatosensory and motor, inferior frontal and parietal, premotor and superior parietal cortices. CRP was also predictive of significantly decreased cortical thickness in 17 areas with the top 10 most significant effects located in effects located in insular and frontal opercular, primary somatosensory, auditory and motor, association auditory, lateral and medial temporal cortices .

In contrast, adult trauma was less strongly predictive of cortical thickness. The unthresholded map of coefficients for the linear regression of AT on cortical thickness was significantly correlated with the map for BMI (*ρ* = *−*0.41, *P_spin_* = 0.0009; Figure 2B). However, its spatial correlation with the effects of CRP was not significant (*ρ* = *−*0.23; *P_spin_* = 0.0815; Figure S9). The relationship between AT and cortical thickness was only significant at 1 cortical area (premotor cortex) after correction for multiple comparisons with FDR *≤* 0.05 (Figure 2 and Figure S8). Cortical effects remained consistent with the use of a coarser parcellation (see SI 1.4).

At the subcortical level, BMI was significantly predictive of increased volume in 6 subcortical areas, including the nucleus accumbens (*β*(*SE*) = 0.0189(0.0068); *t* = 2.78). In contrast, CRP was significantly predictive of decreased volume in three structures: pallidum (*β*(*SE*) = *−*0.0438(0.0068); *t* = *−*6.46), thalamus (*β*(*SE*) = *−*0.0251(0.0068); *t* = *−*3.70), and hippocampus (*β*(*SE*) = *−*0.0187(*−*0.0068); *t* = *−*2.76). Adult trauma was significantly predictive of reduced volume of all subcortical structures tested, including the nucleus accumbens (*β*(*SE*) = *−*0.0339(0.0068); *t* = *−*5.00); for details see Figure 2B and (see ST6, ST5, and ST7).

### Full and sparse path models of adult trauma-, BMI- and CRP-mediated effects of CM on brain structure

We extended the path model of interactions between childhood maltreatment, adult trauma, BMI and CRP to also include MRI measurements of cortical thickness or subcortical volume (Figure 3A&B). This increases the degrees of freedom to estimate path model parameters from six (based on a correlation matrix of 4 variables) to 10 (based on a correlation matrix of 5 variables) at each brain region. As in the prior model (Figure 1), not inclusive of MRI, childhood maltreatment had direct effects on adult trauma and BMI, which both had direct effects on CRP. We fitted and compared two versions of the MRI-inclusive model to test the hypothesis (H3) that childhood maltreatment has indirect effects on adult brain structure.

**Figure 3:**
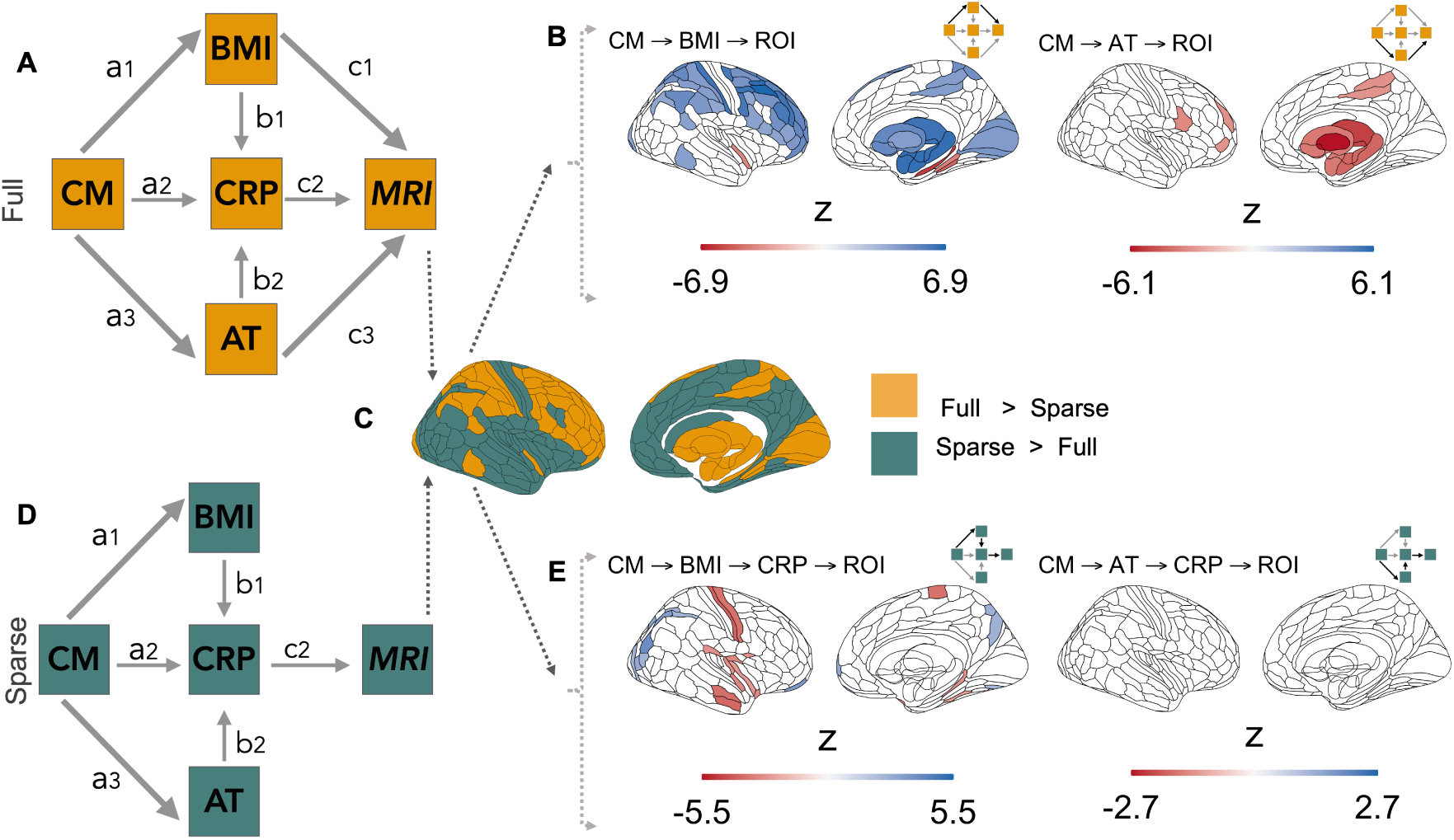
Indirect childhood maltreatment effects on cortical and subcortical structures indexed by two nested path models. **A,D** Diagrams of nested path models of indirect childhood maltreatment effects on brain structure. Full model (**A**): contains direct effects of BMI, CRP and adult trauma on adult brain MRI measurements (paths *c*1*, c*2 and *c*3, respectively); all other paths between non-MRI variables are identical to the model in Figure 1. Sparse model (**D**): contains a direct effect of CRP, only, on adult brain structure; adult trauma and BMI can have indirect effects on brain MRI measurements mediated by their direct effects on CRP. **C** Comparison of full and sparse models: This pair of nested models was evaluated at each of 180 cortical areas defined by the Glasser parcellation [34] and their difference in likelihood-ratio used to determine their relative goodness-of-fit. Yellow areas indicate significantly better fit by the full model at FDR*≤* 0.05; green areas indicate where the sparse model was sufficient to explain local variance. **B, left** Brain map of BMI-mediated indirect effects (Wald *z*) of childhood maltreatment on brain structure, together with a schema highlighting (in bold lines) the paths whose coefficients make up the indirect effect. Blank areas on the cortical and subcortical maps indicate non-significant results (FDR>0.05) or locations where the model was not evaluated. **B, right** Indirect effects of childhood maltreatment on brain structure, mediated by adult trauma. For thresholded and unthresholded maps of all paths evaluated see Figures S11, S12; and for unstandardized coefficients see Figures S13 and S14. **E, left** Childhood maltreatment had indirect effects on adult brain structure that were mediated by a chain of paths from CM *→* BMI *→* CRP. **E, right** There were no significant indirect effects of childhood maltreatment mediated by a chain of paths CM *→* AT *→* CRP. For maps of all thresholded and unthresholded paths see Figures S15 and S16; for unstandardized coefficients Figures S17 and S18. Wald *z*’s represent *β* coefficients standardized by their standard errors.

The two nested models were designated, respectively, full and sparse. The full model (8 parameters) specified that any of the 3 adult variables (AT, BMI or CRP) could have direct effects on brain structure of each region (Figure 3A). The sparse model (6 parameters) specified that only CRP had a direct effect on brain structure, i.e., any effects of adult trauma or BMI on brain structure must be mediated indirectly via their effects on CRP (Figure 3D). Neither model passed the Satorra-Bentler scaled *χ*^2^ test at any region (all *P* <0.05), yet both models had good fit across the entire brain given their CFI, RMSEA, and SRMR scores (see Supplementary Results SI 1.5).

To determine if the full or sparse model represented a better account of the mediated effects of childhood maltreatment on brain structure, we estimated the likelihood ratio for the difference in goodness-of-fit between models at each region (FDR*≤* 0.05). This comparative analysis of performance identified 56 cortical and 6 subcortical structures where the full model performed significantly better in accounting for the data than the sparse model. Whereas the sparse model, where CRP was the ultimate mediator of any indirect effects on brain structure, was a significantly better fit to the data at 124 cortical areas and 1 subcortical structure, the pallidum (Figure 3C). These results suggested that no single model uniformly explained the mediated effects of childhood maltreatment across the brain.

### Indirect effects of childhood maltreatment on adult brain regional structure

To assess the size and significance of hypothetically anticipated indirect effects of childhood maltreatment on brain structure, mediated by the direct effects of childhood maltreatment on adult trauma, BMI and CRP, we estimated the product of path coefficients in each causal chain in both the full and sparse models. We exclusively estimated these coefficients within the set of brain areas where each model was optimally fit (Figure 3C).

In the full model, the product of path coefficients (*a*1 *× c*1) from {CM *→* BMI *→* MRI} was significantly greater than zero within all *N* = 56 cortical and *N* = 6 subcortical areas tested. The strongest indirect effects of childhood maltreatment mediated by adult BMI were concentrated in dorsolateral prefrontal, premotor, and primary somatosensory and motor cortical regions (*z_max_* = 6.89*, β_max_* = 0.00501*, P <* 0.0001); and, subcortically, in the amygdala (*z* = 6.43*, β* = 0.00449*, P <* 0.0001), thalamus (*z* = 6.33*, β* = 0.00437*, P <* 0.0001) and hippocampus (*z* = 6.48*, β* = 0.00455*, P <* 0.0001) (Figure 3B,left).

There was also evidence for an indirect effect of childhood maltreatment mediated by adult trauma on brain structure estimated by the product of path coefficients (*a*3*×c*3) from {CM *→* AT *→* MRI}. This indirect effect passed the threshold for significance in six areas located within premotor, sensorimotor, inferior prefrontal and dorsolateral prefrontal cortical areas (*z_max_* = *−*3.31*, β_max_* = *−*0.00718*, P* = 0.0247), but was significant for all subcortical structures at which the model was evaluated, with the strongest effects in pallidum (*z* = *−*6.84*, β* = *−*0.0133*, P <* 0.0001), thalamus (*z* = *−*5.22*, β* = *−*0.0112*, P* = 0.0002) and nucleus accumbens (*z* = *−*5.06*, β* = *−*0.0109*, P* = 0.0003), (Figure 3B,right).

There were no significant CRP-mediated effects for the product of the coefficients (*a*2 *× c*2) from {CM *→* CRP *→* MRI} (Figure S11). However, effects mediated by the path {CM *→* BMI *→* CRP *→*MRI} were borderline significant for one cortical area (*z* = 3.18*, β* = 0.00080*, P* = 0.0463) and significant for three subcortical structures: pallidum (*z* = *−*6.06*, β* = *−*0.00178*, P <* 0.0001), thalamus (*z* = *−*5.50*, β* = *−*0.00156*, P <* 0.0001) and hippocampus (*z* = *−*5.12*, β* = *−*0.00140*, P <* 0.0001), (Figure S11).

In the sparse model, there was some evidence that indirect effects of childhood maltreatment could be mediated by the longer chain of connections from {CM *→* BMI *→* CRP *→* MRI } although the product of path coefficients was small (*z_max_*= 3.96*, β_max_*= 0.00091*, P* = 0.0009) and only significant in 27 cortical areas (Figure 3E,left). There was no evidence for indirect effects mediated by {CM *→* AT *→* CRP *→* MRI } in the cortex or subcortex (Figure 3E,right).

## Discussion

In this study we addressed three hypotheses concerning pathways linking retrospectively assessed childhood maltreatment to differences in adult brain structure. We tested: (H1) that self-reported experiences of childhood maltreatment partly explain adult inflammation through their effects on adult BMI and trauma; (H2) that adult trauma, BMI and CRP are significantly associated with variation in adult brain structure; and (H3) that childhood maltreatment can have indirect effects on brain structure mediated by its direct effects on adult trauma, BMI and CRP, compounded by the direct effects of these adult variables on the brain. Results largely supported these hypotheses and indicated plausible mechanisms of linkage between early life adversity and adult variables, including brain structure.

### Childhood maltreatment’s influence on BMI, CRP and adult trauma

We first examined how childhood maltreatment was related to adult trauma, BMI and CRP. Path modeling suggested a causal role for childhood maltreatment in higher BMI and higher levels of self-reported adult trauma. We also found that, although childhood maltreatment does not have direct effects on adult CRP, or near-negligible effects only discoverable in the much larger N=116,887 UKB cohort, it can have pro-inflammatory effects indirectly via its direct effects on BMI and adult trauma. Higher blood concentrations of pro-inflammatory signalling molecules have been well documented in adults exposed to maltreatment in childhood [7]. However, there are not yet sufficient mechanistic explanations for this association [83]. Our results indicate that adult obesity following childhood maltreatment could be a driver of systemic inflammation in adulthood, and that this pro-inflammatory state can be exacerbated by re-exposure to traumatic stressors in adulthood.

Childhood maltreatment is linked to a host of atypical cognitive processes underlying maladaptive behaviors that likely mediate the increased risks for obesity and traumatic experiences in adulthood [64, 107]. Such atypical cognitive processes include heightened social threat monitoring [19, 80, 65, 68]; impaired learning and cognitive control, i.e., executive dysfunction, and increased emotional reactivity [40, 59, 24]; and reduced reward sensitivity [9, 62, 42]. These differences in cognition and behavior are thought to arise from reduced volumes and altered connectivity in salience and prefrontal-amygdalar, fronto-striatal and fronto-parietal networks; circuits involved in threat-detection, reward processing, and cognitive control-respectively [67].

The cognitive impact of maltreatment may act in tandem with other early life stress-induced physiological alterations and give rise to a so-called “thrifty phenotype” that leads to increased BMI [20]. This phenotype is distinguished by greater energy intake and storage and/or decreased energy expenditure [20]. In this context, individuals are driven to the consumption of calorie-dense foods through reduced reward sensitivity and executive dysfunction [70, 107], emotional dysregulation induced use of food as a soothing mechanism [24], and neuroendocrine-mediated deviations in appetite [3]. Conversely, lower energy expenditure is often regarded as a downstream consequence of obesity primarily due to excessive food intake, where inflammation-associated fatigue, and increased physical mass convergently reduce daily activity levels [3, 72].

Higher rates of adult trauma may also be caused by the cognitive sequelae of childhood maltreatment. For instance, social cognition in maltreated adolescents is best explained by the severity of their exposure to these experiences[18]. Adolescents who have experienced maltreatment as children are seen as less socially competent by their peers and teachers [51, 61]; and, as adults, they have more dysfunctional interpersonal relationships [46]. Heightened monitoring of social threats, poor cognitive control over emotions, and lower responsivity to social rewards may drive difficulties in social functioning by decreasing an individual’s ability to successfully negotiate complex social environments, and by increasing life-time vulnerability to stressors and material precariousness [63].

*How can BMI and adult trauma influence inflammation?* Body mass index scales linearly with mass of visceral adipose tissue, which is responsible for generating and regulating immune signaling molecules, including IL6 and other pro-inflammatory cytokines [10]. In human observational studies and animal experiments, greater fat mass is robustly positively correlated with increased blood cytokine concentrations [44], making obesity a cause of systemic inflammation. The pathway between self-reported adult trauma and heightened CRP levels is less obvious, though the neuroendocrine system is a candidate mechanism for linkage. The neuroendocrine system is involved in the regulation of physiological responses to stress, and eventual recovery of homeostasis, via the hypothalamic-pituitary-adrenal (HPA) axis which releases and regulates cortisol, an anti-inflammatory glucocorticoid hormone [102, 91]. Chronic trauma or stress leads to sustained cortisol secretion and ultimately dysregulation of the glucorticoidreceptor leading to failure to downregulate inflammatory signalling pathways despite high circulating levels of cortisol [56, 45].

### CRP-, BMI- and trauma-related variation in brain structure

We found that CRP, BMI and adult trauma were all independently related to brain structure. Adult trauma had relatively minor effects on cortical thickness but was strongly associated with volume reductions of all subcortical structures tested. In contrast, BMI and CRP both had widespread and anatomically convergent effects on cortical structure, with divergent and more localised effects on subcortical volumes (Fig. 2). This pattern of results suggest that cortical structure may be sensitive to maltreatment-induced dysregulation of metabolic and/or immune systems; whereas adult trauma impacts brain structure by different biological pathways.

Lower subcortical volumes in relationship to adult trauma have been previously reported [108, 5]. Limbic structures are rich in GR receptors and expression density of these receptors can be down-regulated by chronic cortisol secretion [77], subsequently inducing deleterious changes in neuronal plasticity and integrity [66]. This may constitute an independent pathway by which subcortical brain structure is modulated by trauma in adulthood.

The strongest signal for immuno-metabolic effects of CRP and/or BMI was increased cortical thickness in prefrontal, frontal and somatosensory and premotor cortical areas. We also observed decreased thickness of insular, frontal opercular and primary somatosensory cortices in association with increased BMI and CRP, and decreased volume of medial temporal cortex associated specifically with higher BMI. Global increases in CT have been associated with increased BMI [86], although this is the first-time that comparable BMI associations have been reported for regional thickness of multiple cortical areas. Nonetheless, our overall pattern of effects is echoed in previous studies of grey matter structure in relation to increasing BMI. We replicate the reductions in thickness of temporal and medial-frontal regions [73, 69, 101, 27] and even though the trend across prior volumetric studies is towards cortical volume reductions [38, 15, 39], some studies have reported increased volume of occipital and frontal cortical regions associated with BMI [43, 76, 98]. Prior trends in association of BMI with subcortical volumes are mixed [39, 30, 50, 73], although several studies find increases in nucleus accumbens [30], hypothalamus [43, 8] amygdala and hippocampus [73, 106] with greater BMI.

Heterogeneity of results across the published literature might emerge due to methodological variability. For instance, positive associations between volume and BMI might have been previously obscured by sample size and composition: non-significant volumetric increases in hippocampal [104], and occipito-frontal regions [82] have been reported in small-sample (*N* <100) studies, with more widespread cortical and subcortical effects being apparent with larger (*N* >1000) samples [98, 73] or extensive exclusion criteria for cardiovascular and neuropsychiatric diseases [43, 76]. This suggests that the full pattern of medial frontal and temporal cortical decreases, and frontal and occipital cortical increases, associated with obesity might have been previously attenuated by lack of statistical power.

The overall pattern of immuno-metabolic effects on brain structure could be driven by systemic circulation of pro-inflammatory cytokines that can trigger astrocyte-dependent neuroinflammatory processes apparent as both increases and decreases in grey matter thickness and volume [72]. Whilst decreases in MRI estimates of thickness may arise due to neuronal damage [72], increases can be a signal of both deleterious and neuroplastic effects [90]. Greater hippocampal volumes are observable in animal models of both exercise and high fat diets; with the later being exclusively driven by histologically-confirmed neurogenesis [90]. A putative mechanism for volumetric increases in high fat diets is an inflammation-driven increase in extracellular free-water content, which has been shown to scale linearly with CRP across cortical regions of the default-mode network in humans [52], and across most of the brain with increasing BMI [93, 54].

### Pathways from childhood maltreatment to variation in adult brain structure

Finally, we found that the effects of BMI, CRP and adult trauma on brain structure could be partially attributed to childhood maltreatment’s prior effects on these adult variables. Our comparison of full vs sparse path models suggested that temporo-parietal reductions in thickness are best explained by a causal chain of the form {CM *→* BMI *→* CRP *→* MRI } whereas frontal, occipital and subcortical increases in grey matter are better explained by a direct effect of BMI that is not contingent on the immune state indexed by CRP.

A large body of literature links childhood maltreatment to adult brain structure [71]. Many of these alterations are already observable in childhood, subserving the atypical cognition associated with maltreatment [67] and likely giving rise to obesity and psychosocial dysfunction [67, 3, 63]. However, previous highly powered studies in older adults have shown that wide-spread alterations in brain structure are most apparent when considering the interaction between maltreatment and age [28]. This suggests that physiological factors exerting progressive influence on brain development over time must be fundamental to our understanding of how maltreatment becomes biologically embedded and increases lifetime risk of psychopathology [45]. Our models were able to provide additional insight by showing that variation in adult brain structure after maltreatment is attributable to two distinct pathways of action: an immuno-metabolic path, linked to wide-spread differences in cortical thickness and subcortical increases in volume, and a distinct, putatively neuroendocrine pathway linked to reductions in limbic volumes. These alterations in brain structure indirectly caused by distinct aspects of the physiological response to adversity therefore represent a plausible set of mechanisms by which childhood maltreatment can have sustained effects on risk for psychopathology.

### Limitations

There are several limitations to this study. First, path models with cross-sectional data do not allow us to draw conclusions on causality. Therefore, inferences on the directionality of relationships between our contemporaneous variables (BMI, CRP, AT) are limited. However, the extent of model-to-data agreement allows us to assess whether the hypothesized relationships are in good agreement with observed correlations between variables [2]. A further limitation of our methods is the use of retrospective self-report as an index of childhood maltreatment. Meta-analyses have shown that retrospective, as opposed to prospective, measures are more sensitive to recall bias due to the effects of adult psychopathology [4]. Therefore the neurobiological profile of people exposed to childhood malreatment assessed by retrospective self-report might be biased towards individuals with a past or current history of mental ill-health. Although we control for adult socioeconomic status (SES), which correlated with childhood SES [97] a more direct measure of this variable would be more desirable in this context, as childhood SES which has been shown to induce changes in cognitive function that exist alongside other risks associated with childhood maltreatment and likely enhance their impact on the individual [22, 99]. Attempts to replicate and extend the findings of this study would benefit from including variables that more directly index the HPA-axis, which was not possible here due to the lack of cortisol measures in the UK Biobank.

### Conclusions

Here we show that childhood maltreatment can exert long term indirect influences on adult brain structure through the physiological response to chronic adversity in immuno-metabolic and psychosocial systems as indexed by BMI, CRP and adult trauma. We therefore provide a mechanistic explanation as to how psychopatholoy risks may be sustained across the lifetime after childhood maltreatment.

## Methods

### Data and sample

Data were obtained from the UK Biobank (UKB) (study ref no. 20904) which is a cohort of approximately 500,000 participants (ages 39 to 73), recruited from the United Kingdom population between 2006 and 2010, who completed extensive biological phenotyping and were additionally assessed with a follow-up online mental health and behavior questionnaire between 2012 and 2013 [96]. Ethical approval for the use of these data was provided by the UKB research ethics committee, and the Human Biology Research Ethics Committee, University of Cambridge (Cambridge, UK). Principal analyses focused on a subset of participants (N*∼*40,000) for whom MRI data of the brain were acquired and both T1- and T2-weighted images passed quality control (QC) (UKB MRI sample).

Eligibility criteria for this sample are detailed in Supplemental Information SI 1.1. UK Biobank participants who did not have MRI data but who otherwise met eligibility criteria formed an internal replication sample (UKB sample; N=21,738) for non-MRI analyses; see **Supplementary Information (SI) Methods** SI 1.1.

### Immune, metabolic and psychosocial phenotypes

#### C-reactive protein (CRP)

C-reactive protein was measured in blood serum by high sensitivity immuno-turbidimetric assays (units: mg/L), on a Beckman Coulter AU5800.

#### Body mass index (BMI)

Body mass index (*kg*/*m*^2^) was derived from weight (kg) and height (m) measurements with a Tanita BC-418 body composition analyser and a Seca 202 measuring rod respectively.

#### Childhood maltreatment (CM)

Self reports of childhood maltreatment were measured by the childhood trauma screener (CTS) [36]. The CTS assesses emotional, physical and sexual abuse, as well as emotional and physical neglect in childhood [32]; for details see SI 1.2.1.

#### Adult trauma (AT)

Adult trauma scores were defined as the sum total of scores on 5 items from the UKB mental health questionnaire that assessed adult experiences corresponding to the early life experiences measured by the CTS; for details see SI 1.2.2. These items indexed emotional, physical and sexual abuse in interpersonal relationships as well as poor closeness of relationships and financial security after age 16 [23].

### Brain MRI phenotypes

#### MRI data: pre-processing, quality control

Details for MRI acquisition protocols in UKB can be found in SI 1.2.2. Downloaded 3D-MPRAGE T1-weighted images were pre-processed according to the Human Connectome Project (HCP) minimal Freesufer pipeline [35]. This pipeline includes artifact removal, pial and cortical surface generation, cross-modal registration, and standard-space alignment. T2-weighted images were used to derive more accurate surface representations [33, 35]. Subjects without T2-w scans whose images have been processed with the HCP pipeline have been shown to have biased morphometric measures [95] and therefore participants without T2-w data were excluded from this analysis. Freesurfer’s reconstruction quality was quantified with the Euler index [87]. Each image had the cortex parcellated according to the Glasser brain atlas (360 cortical regions), from which the macro-structural measures of cortical thickness (CT) were estimated [34]. Volumetric partitions of 14 subcortical structures were also defined by Freesurfer [26].

A regional measurement in an individual scan, i.e., *CT_i,j_* the cortical thickness of the *i*th region in the *j*th participant, was excluded from analysis if it was more than 5 median absolute deviations (MAD) from the regional median, i.e., *CT_i,j_ ±*5*MAD*. Both volume and CT measures were corrected for sex, age at the time of scanning, sex*×*age interaction, and the Townsend deprivation index as a proxy measure of socio-economic status. We also corrected for batch effects due to scanning at multiple UKB imaging centres [25]; position of the head and radiofrequency receiver coil; and frame-wise displacement (FD) - a frame-to-frame index of head motion derived from functional MRI data that can serve as a proxy measure of motion-induced bias during structural MRI scanning in the same session [84, 92]; for more detail see SI 1.2.2.

Finally, bilaterally homologous MRI measures were averaged for each participant, resulting in data on CT at each of 180 symmetrized cortical areas and volume of 7 symmetrized subcortical structures, see SI 1.2.5.

### Statistical analyses

All analyses were conducted in RStudio (R version 4.2.2)[81]; see SI 1.2.6.

#### Distribution and nuisance correction of non-MRI phenotypes

All non-MRI phenotypes (CRP, BMI, CM and AT) displayed a high distributional skewness (Figure S2A). To correct for this, natural scale values were log-transformed (Figure S2B) and used as the basis for all analyses. Additionally, all measures were corrected (i.e. nuisance regressed) for sex, age at the time of baseline visit, sex*×*age interaction and the Townsend deprivation index (socioeconomic status. For results of these regressions see ST1 and ST2. Correlations between all variables were estimated with Spearman’s method.

#### Linear models correction for spatial auto-correlation

We used linear regression to address the relationship between CRP, BMI, and AT and brain structure (H2). When examining the correlation between the resulting spatial maps, spin tests were used to derive *P* -values corrected for spatial autocorrelation [29].

#### Path analysis

A path model depicts a theoretical set of directed relationships between correlated variables, which then serves as the basis for estimation of path coefficients from the correlation matrix. Path analysis can be regarded as an extension of multiple regression that allows for statistical inference on hypothetically directed relationships [53]. Note that the technical capacity to estimate directed or asymmetrical paths between variables does not double as a tool for providing evidence of causality when any two variables are contemporaneously measured. In such cases, a good-fitting path model can serve as a useful indicator of agreement between theory and data [11], but it does not substantiate an underlying causal mechanism.

Path models were fitted to maximise the likelihood of estimated path coefficients using the ”lavaan” package in R [89]. To obtain reliable statistics despite remaining non-normalities in the log-transformed data, we implemented robust maximum likelihood estimators with Huber-White standard errors (MLR) which are asymptotically equivalent to a Yuan-Bentler test statistic [89]. The maximum number of path coefficients, *p*, that can be estimated is the degrees of freedom *df* = *m*^2^ *− m*/2, where *m* is the number of variables in the correlation matrix. For non-saturated path models, with *p ≤ df*, we measured model fit to the data with the following measures: the Santorra-Bentler scaled *χ*^2^ test, the comparative fit index (CFI), the root mean square error of approximation (RMSEA) with its confidence interval, and the standardized root mean squared residuals (SRMR) [94]. We report the robust variants of these measures due to the use of an MLR estimator [12]. We evaluated model goodness-of-fit by the following criteria: CFI, acceptable fit 0.95-0.97, good fit >0.97; SRMR, acceptable fit 0.05–0.10, good fit <0.05; and RMSEA, acceptable fit 0.05-0.08, good fit <0.05. For the models fit to the UKB MRI sample, all paths (direct or indirect) were tested for significance using the false discovery rate (FDR) at 5% to correct for multiple comparisons, where the number of tests considered was equal to the number of anatomical regions tested (*N* = 187) when constructing a symmetrized whole brain map.

Direct effects between variables are represented by a single path, e.g., *A → B*, in the model and their strength is estimated by a single path coefficient, e.g., *a*. Indirect effects between variables are represented by a chain of paths, e.g., *A → B*, *B → C* for the indirect effect of *A* on *C*, and a corresponding series of path coefficients, e.g., *a, b*. The product of the path coefficients *a × b* is a measure of the size of the indirect effect, and is Normally distributed for large *N* s curtailing the need for non-parametric methods of statistical inference [1], see SI 1.2.5.

#### Model contrasts

The MLR fit functions used to estimate path coefficients yield a *t*-statistic which is the basis for comparison between nested models. For the UKB MRI sample, we fitted two nested models and contrasted their performance at each region with a likelihood-ratio test where the difference in *t*-statistics between the two models under the null hypothesis follows a *χ*^2^ distribution with *k* degrees of freedom where *k* = *p*_1_ *− p*_2_ and *p*_1_*, p*_2_ denote the number of path coefficents estimated in models 1 and 2, respectively. Likelihood ratio (LR) statistics were tested for statistical significance with FDR = 5% to correct for multiple comparisons and significant LR testsindicates that the data was best explained by the more complex model, with a larger number of path coefficients, at that region.

## Data Availability

All data used in the present study are available upon successful application at https://www.ukbiobank.ac.uk/

## Acknowledgements

We thank the participants of the UK BIOBANK who provided their data along with the researchers inolved in data collection and assembly. This work was funded by: an MPhil studentship from Girton College (S.C.O), University of Cambridge; a PhD studentship from Darwin College, University of Cambridge (S.C.O); NIH Research Senior Investigator award (to E.T.B.); MQ: Transforming Mental Health grant MQF17_24 (to P.E.V.); the NIHR Cambridge Biomedical Research Centre (NIHR203312) and the NIHR Applied Research Collaboration East of England. The views expressed are those of the author(s) and not necessarily those of the NIHR or the Department of Health and Social Care.

