## Supplementary materials for "Childhood maltreatment influences adult brain structure through its effects on immune, metabolic and psychosocial factors"

### Supplementary Methods

#### SI 1.1 Sample selection

The UK BIOBANK provided participant data for this study. Principal analyses focused on a subset of participants invited for a multi-modal MRI follow-up measurement for whom, at the time of analysis, N=40,680 had images of the brain available for download. Exclusion criteria consisted of the following: (1) imaging data was deemed low quality (see SI ”MRI post-processing and quality control”); (2) data on socieconomic status (Townsend Deprivation Index) was missing; (3) C-reactive protein assays were missing; (4) Body Mass Index data was missing; (5) any of the mental health questionnaire items relevant to this study were unanswered. Participants who met criteria 2-5 but for whom MR images were not acquired or were deemed low quality were selected as a replication sample for non-imaging analyses. Figure S1 illustrates our sample selection protocol showing that the imaging sample has N=21,738 subjects and the non-imaging sample has N=116,887.

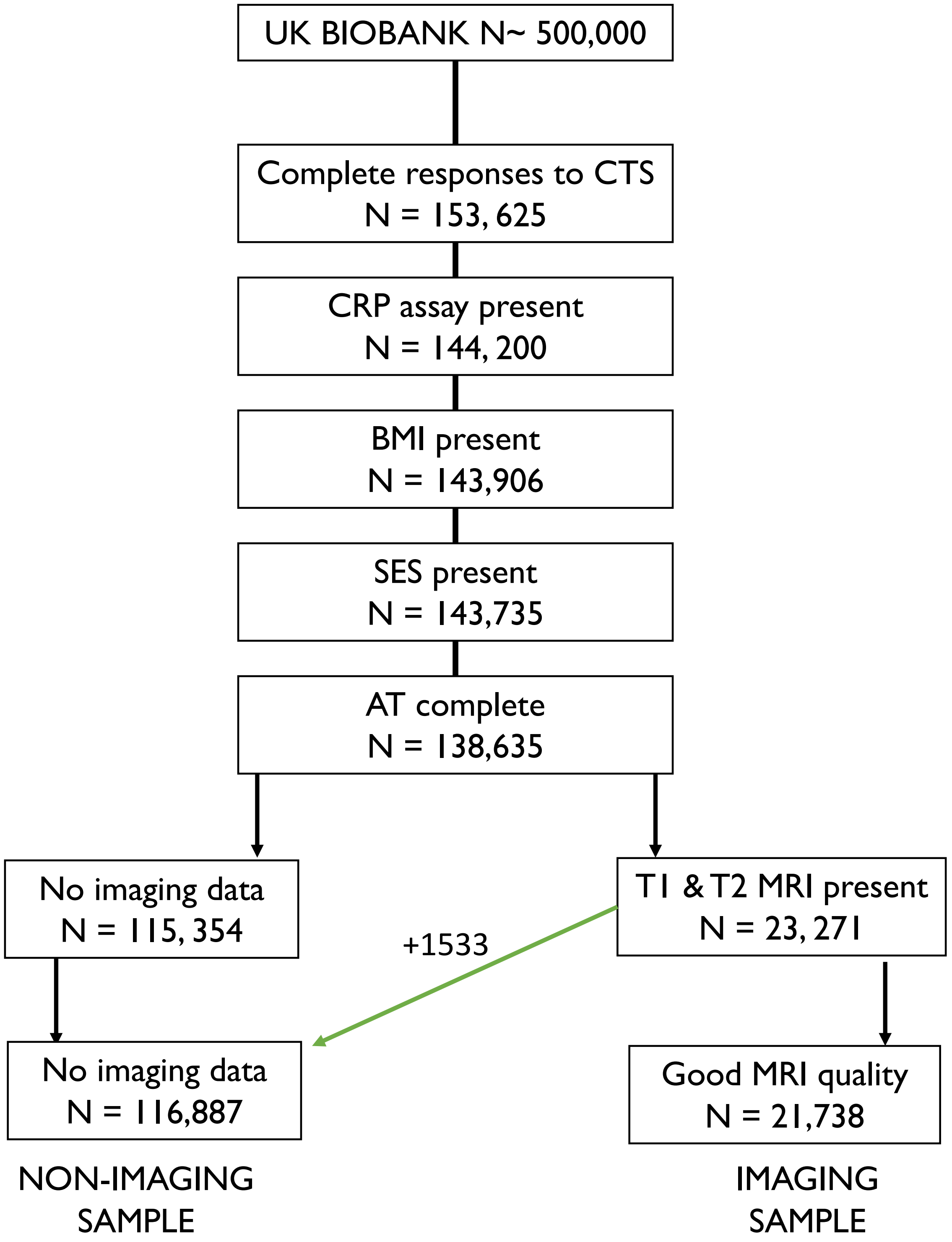

**Figure S1:** **Sample selection algorithm.** CTS = childhood trauma questionnaire; BMI = body mass index; SES = socieconomic status as indexed by the Townsend Deprivation Index; AT = adult trauma questionnaire; MRI = magnetic resonance imaging.

#### SI 1.2 Immune, metabolic and psychosocial phenotypes

##### SI 1.2.1 Childhood maltreatment (CM)

Items corresponding to the childhood trauma screener (CTS) [10, 14] were extracted from the online mental health questionnaire developed by the UK BIOBANK Mental Health steering group [6] and a total "Childhoood Maltreatment" sumscore was built with them. The CTS is a shortened version of the commonly implemented childhood trauma questionnaire (CTQ) [4].

##### SI 1.2.2 Adult Trauma (AT)

Adult trauma scores were derived from relevant items of the UK BIBANK online mental health questionnaire [6]. The questionnaire items from which the total sum scores were constructed are the following: (1) I have been in a confiding relationship (reversed); (2) A partner or ex-partner deliberately hit me or used violence in any other way; (3) A partner or ex-partner repeatedly belittled me to the extent that I felt worthless; (4) A partner of ex-partner sexually interfered with me, or forced me to have sex against my wishes; (5) There was money to pay the rent or mortgage when I needed it (reversed). These items were responses to the prompt "since I was sixteen", indicating that subjects could report events relevant to the items that took place after childhood and early adolescence. Responses were given by a 0-4 Likert scale with 0 = "Never true" and 4 = "very often true." Individuals responding "prefer not to answer" were excluded. Positive items were reverse coded, with higher total scores indicating greater trauma.

##### Phenotype log transform

Due to the high distributional skewness (Figure S2) of childhood trauma, adult trauma, BMI and CRP these variables were log-transformed; transformed values then formed the basis of all analyses.

#### MRI acquisition and processing

Structural magnetic resonance images of the whole brain were acquired on Siemens Skyra 3T scanners with 32-channel head coils. Acquisition took place with three identical scanners at three different dedicated imaging centers [16]. Reconstruction of images from k-space, and bias field correction took place in the scanner with standard Siemens software, without applying gradient distortion correction [2]. Further details on the UK BIOBANK's imaging protocol can be found in:

- http://biobank.ctsu.ox.ac.uk/crystal/refer.cgi?id=2367
- http://biobank.ctsu.ox.ac.uk/crystal/refer.cgi?id=1977

Next, 3D MPRAGE T1-weighted scanner-pre-processed images were then pre-processed according to the Human Connectome Project (HCP) minimal Freesurfer pipeline [13]. Processing included, artifact removal, pial and cortical surface generation, cross-modal registration, and alignment to standard space. When available, T2-w FLAIR images were used in order to derive more accurate surface representations [13, 11]. Euler indices were derived as a proxy measure of Freesurfer's reconstruction quality [21]. The cortex was anatomically segmented according to the Glasser brain atlas into a total of 360 regions [12], and cortical thickness (CT) estimates were derived for each.

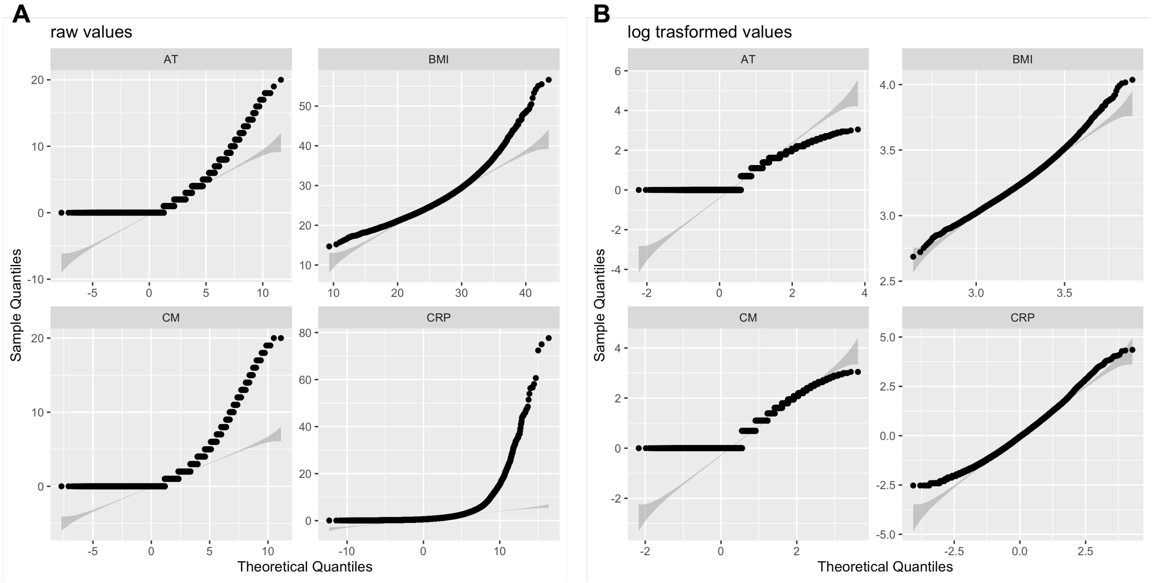

**Figure S2: QQ plots of immune, metabolic and psychosocial phenotypes. A.** Raw values as supplied by UK BIOBANK (CRP and BMI) or as sumscores of UK BIOBANK questionnaires (childhood maltreatment (CM) and adult trauma (AT). **B.** $log_{10}$ transformed phenotype values

Volumetric partitions of the subcortex were yielded from the ASEG atlas[9]. We only make use of the resulting grey matter structures in analyses: thalamus, caudate, putamen, pallidum, hippocampus, amygdala and accumbens.

##### SI 1.2.3 MRI post-processing and quality control

Cortical thickness estimates have been shown to be consistently biased if these were estimated without T2-w FLAIR input [15, 1]. Therefore, subjects without these data were excluded from imaging analyses. In this sample, those without a T2-w image had consistently lower mean cortical thickness values [26]. This is in line with previous observations that absence of FLAIR images for pre-processing is an important source of confounds in UK BIOBANK [1]. Additionally, and prior to nuisance correction, we excluded a given individual's ROI from analyses if it was a CT or volume outlier with a deviation 5 times the median absolute deviation ($\mp$ 5 MAD).

##### SI 1.2.4 Imaging nuisance variables details

Both CT and volume estimates were corrected for imaging confounds. The basic confounds considered were sex, age at scanning, the sex and age interaction, and the Townsend deprivation index - our proxy measure of socioeconomic status (SES). Additional parameters included BIOBANK imaging centre effects [8], Freesurfer's Euler index –a reliable measure of data quality [21]-, head position in the scanner, and framewise displacement (FD). FD is an index of frame-to-frame head motion derived from functional MRI data. Head motion during structural image acquisition is capable of introducing artifacts that bias CT and volume estimates [20]. FD has been shown to have high within- subject stability across different sessions of fMRI acquisition, and it is therefore possible to assume that it provides a sensible estimate of subject motion during structural MRI acquisition [23]. Finally, positioning of both the head and radio-frequency receiver coil were considered due to their tendency to vary across participants, potentially introducing bias. Head position was characterized through x,y,z coordinates, plus z- positioning of the coil.

##### SI 1.2.5 Bilateral atlas construction

We ran regression models for AT, BMI, CM and CRP independently as predictors of cortical thickness or subcortical volume across all 374 ROIs in the Glasser cortical and aseg subcortical atlases. Symmetrical effects can be observed for the cortex and subcortex across all variables (fig.S3A&B) and these are shown to be highly correlated (fig.S3C). Due to these observations we averaged region-wise left and right values of CT or volume, for the cortex and the subcortex respectively, yielding a new atlas of 187 ROIs (180 cortex, 7 subcortex).

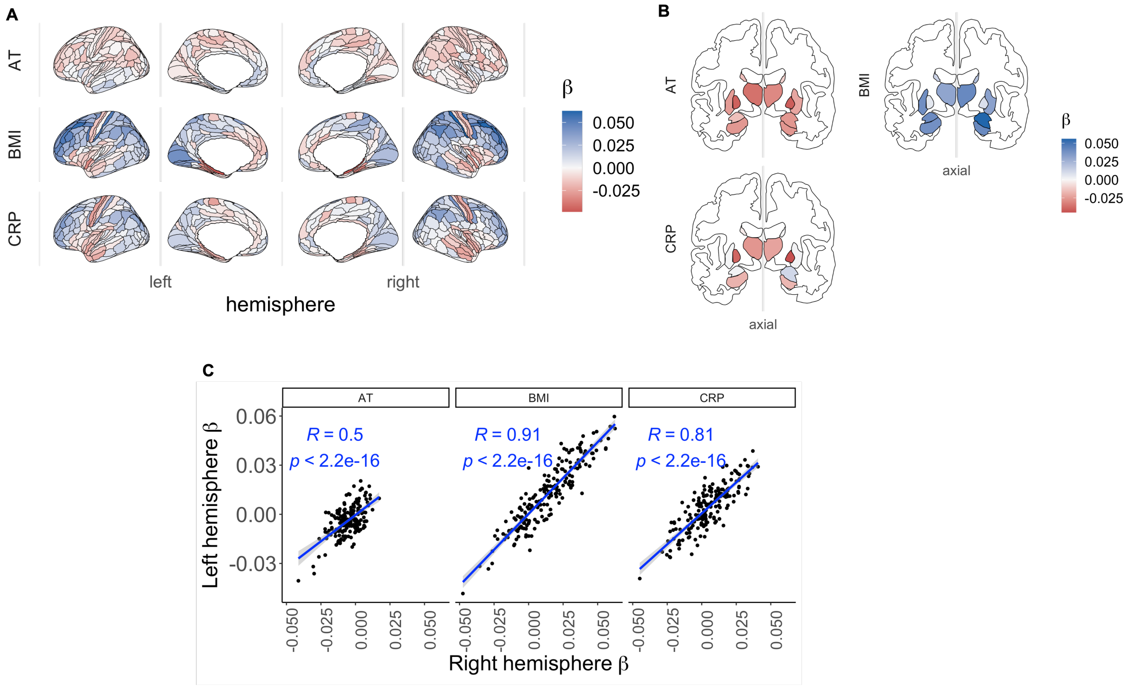

**Figure S3: Symmetry of brain structural effects of AT, CRP, and BMI across hemispheres**. **A**. Independent linear effects for AT, CRP and BMI on cortical thickness for all non-lateralized 360 cortical areas. $\beta$ unstandardized beta coefficients. **B**. Independent linear effects for AT, CRP and BMI on subcortical volumes for all non-lateralized 14 subcortical regions **C**. Scatterplots of unstandardized effects ($\beta$) correlations (Pearson, $\rho$) between left and right hemispheres.

##### Estimation and inference of indirect path model effects

The product of the path coefficients $a \times b$ is not normally distributed for small $N$s [5]. In such cases, inference often relies on the construction of non-parametric sampling distributions for the product of coefficients through either resampling (e.g., bootstrapping) or simulation (e.g., Monte Carlo) procedures [7, 19]. However, for extremely large $N$s, such as those in our UKB MRI sample, the joint distribution of the products approaches normality [3, 18]. Significance of the product of coefficients can then be assessed with the Delta method, lavaan's default choice for statistical inference, which solely relies on the variance of coefficients being multiplied to obtain the standard error and associated t-statistics of their joint distribution [25, 22]. Due the the large size of both of our samples we therefore relied on the Delta method for statistical inference in our indirect paths.

However, for data that has been scaled before fitting the models lavaan is unable to correctly estimate the p-values of extremely small coefficients. For the indirect effects where this is the case, we simply report the unstandardised coefficients and automatically treat these as non-significant.

##### SI 1.2.6 Software

All analyses were conducted in RStudio version 2022.12.0+353 running R version 4.2.2 using the packages: lavaan [22], dplyr [29], and tidyverse [30]. Plots were constructed with the package ggplot2 [28] and brain surface plots were created with ggseg and ggsegglasser [17, 17]

### Supplementary Results

#### SI 1.3 Relationships between childhood maltreatment, adult trauma, BMI and CRP in the larger UKB sample

We repeated analyses verifying our first hypothesis (H1) which states that BMI and AT mediate the relationship between CM and CRP. These replication analyses were conducted on on all subjects of the UK BIOBANK who met our inclusion criteria but had no brain imaging data, or whose imaging data was of poor quality. H1 results were replicated on this sample except for the direct relationship between CRP and CM. In this sample, which is at least five times as large ($N$ = 116,887), there was a direct of effect of childhood maltreatment on CRP ($CM \rightarrow CRP, z = 3.431, P < 0.001$). See Figure S4B and Table ST4 for details. Model fit in this sample was good: $SRMR = 0.010; CFI = 0.997; RMSEA = 0.031$.

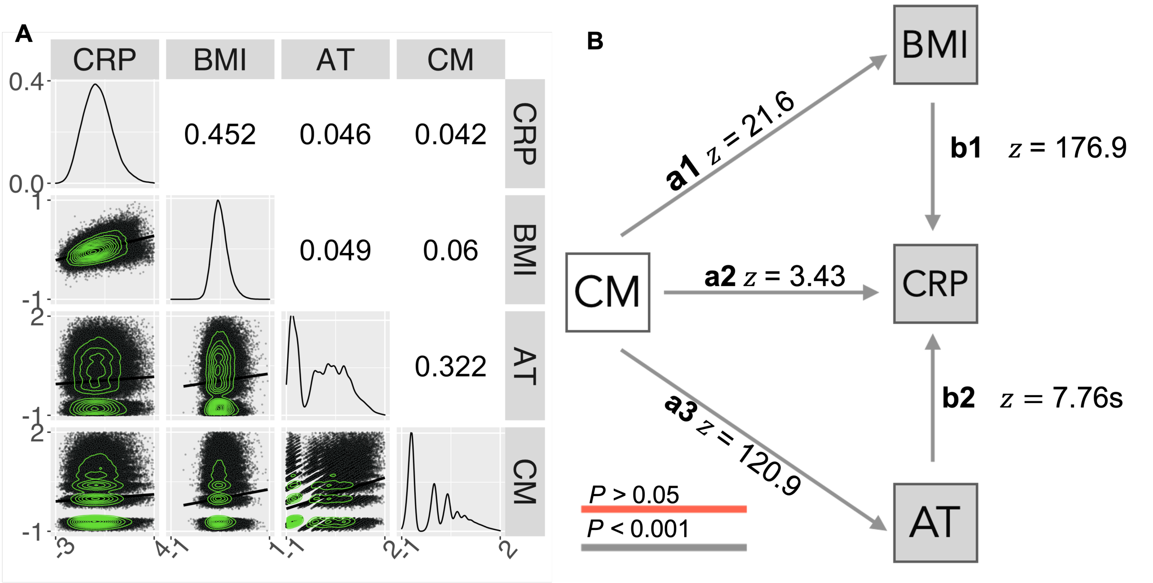

**Figure S4: Replication of relationships between childhood maltreatment, adult trauma, BMI and CRP in the larger UKB sample**. **A**. Correlation matrix representing pair-wise Spearman's correlations (upper triangle) and scatterplots of the relationships between each pair of variables, with solid lines indicating fitted linear regression models (lower triangle). The diagonal represents the probability density function for each variable. All correlations were significantly greater than zero, with FDR $\leq 0.05$. **B.** Path diagram representing direct effects of retrospectively ascertained CM (white) on the contemporaneously measured adult variables, AT, CRP, and BMI (grey). Standardized path coefficients are given as Wald ($z$) statistics

#### SI 1.4 Replication of effects of adult trauma, BMI and CRP on cortical thickness with a coarser parcellation.

We examined whether our results examining the effects of AT, BMI and CRP on CT remained consistent when using a coarser parcellation, or brain map, with a smaller number of areas. We leveraged the fact that the Glasser brain atlas groups each of its 180 areas into 22 distinct and spatially adjacent brain regions [12]. We then averaged cortical thickness of each set of areas within a given region, yielding 22 regional estimates of cortical thickness. This approach has been followed elsewhere [27]. Next, Adult trauma, BMI and CRP were each treated separately as independent variables in three different linear regression models with cortical thickness as the dependent variable at each of the 22 *new* cortical areas. For all variables, the resulting brain maps remained qualitatively consistent with the main results relying on the more granular 180-area parcellation, see Figure S5.

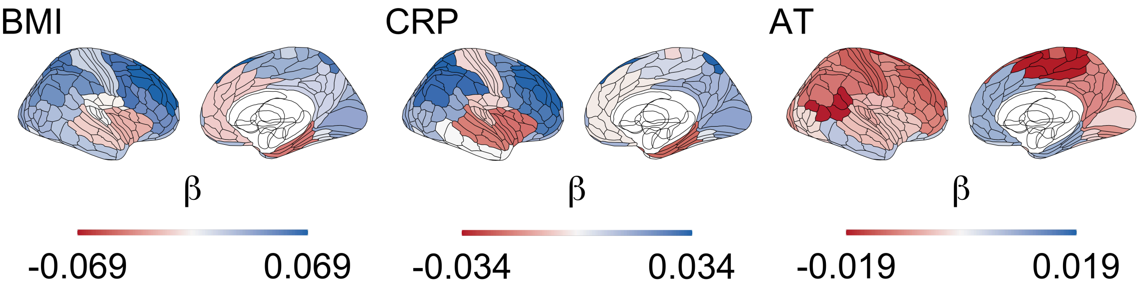

**Figure S5: Unthresholded brain maps of independent relationships between adult trauma (AT), BMI, or CRP with coarser parcellation of the Glasser atlas**. Before obtaining empirical results, each area was grouped within one of Glasser's brain regions (or groupings) [12] and the average within-region cortical thickness was computed. Lines delineating the original brain regions are left as visual reference on the brain maps. $\beta$ = unstandardised regression coefficients.

#### SI 1.5 Complementary goodness of fit assessments

Following the convention in path modelling for evaluating goodness of fit, we supplement the Satorra-Betler $\chi^2$ tests with the following additional indices: Comparative Fit Index (CFI), the root mean square error of approximation (RMSEA) with its confidence interval, and the standardized root mean squared residuals (SRMR) [24]. We evaluated each model at $n = 180$ cortical areas and $n = 7$ sub-cortical structures and derived their goodness of fit measures. Results for the full and sparse models can be seen in figures S6 and S7 respectively. Neither model passed the Satorra-Bentler scaled $\chi^2$ test at any region (all $P < 0.05$).Note that this test is extremely sensitive to small discrepancies between the observed correlation matrix to which the model is fit and the matrix *implied* by a given path model. Therefore, significant p-values (which signal poor data and model agreement) are often expected for large $N$s and fitness is evaluated jointly with other measures [24]. In this case both models can be considered to have good fit across the entire brain given their good CFI, RMSEA, and SRMR scores (see figures S6 and S7 ).

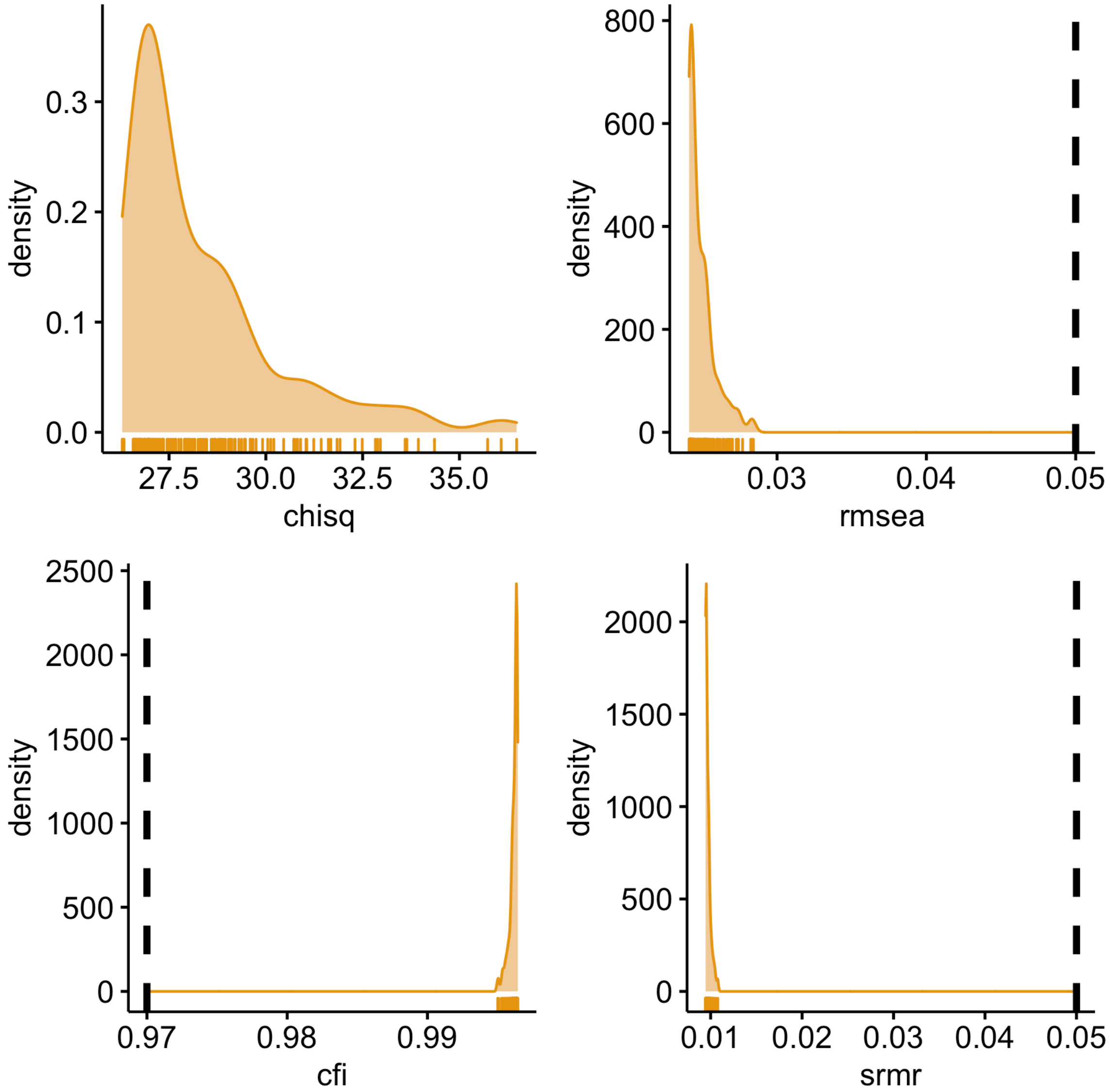

**Figure S6: Full model goodness-of-fit indices density plots for the whole brain**. Dashed lines indicate good fit thresholds, where rmsea and srmr are deemed good if <0.05, and cfi >97.

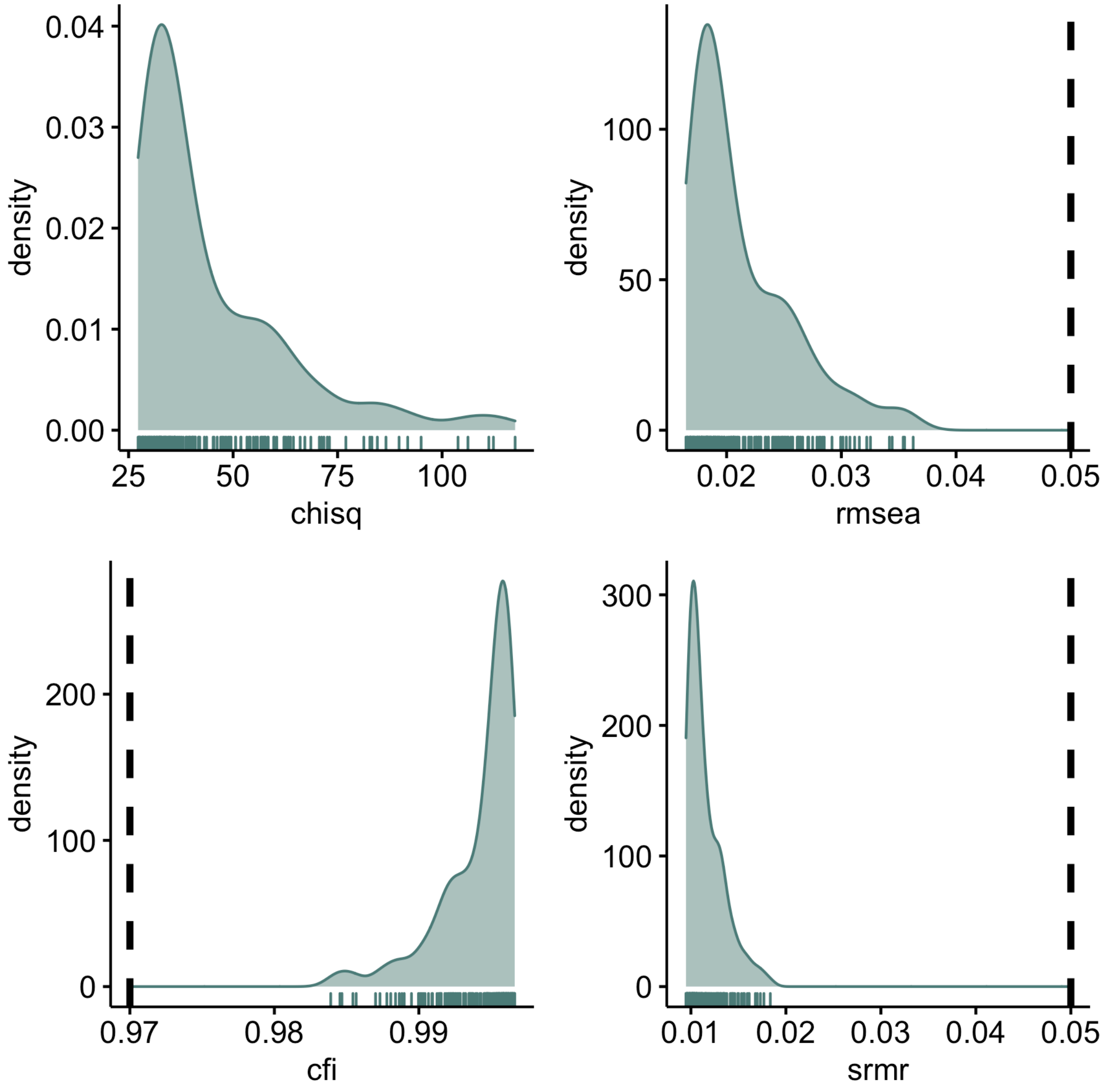

**Figure S7:** **Sparse model goodness-of-fit indices density plots for the whole brain**. Dashed lines indicate good fit thresholds, where rmsea and srmr are deemed good if <0.05, and cfi >97.

### Supplementary Figures

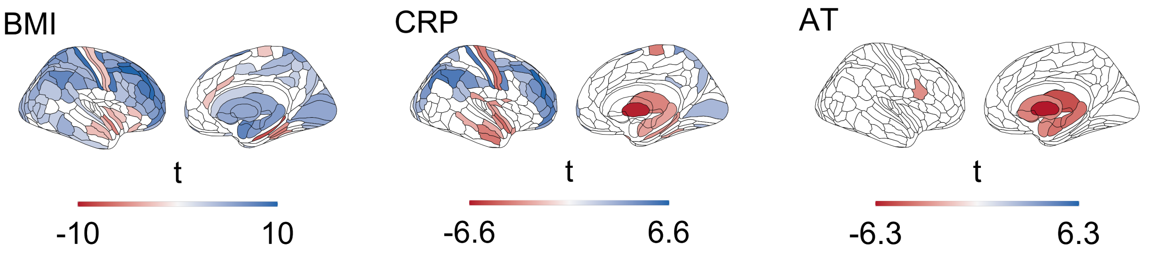

**Figure S8: Thresholded brain maps of independent linear relationships between adult trauma (AT), CRP or BMI and cortical thickness and subcortical volume.** Each map shows the anatomical distribution of thresholded effects at FDR $\leq 0.05$ effects ($t$ values)

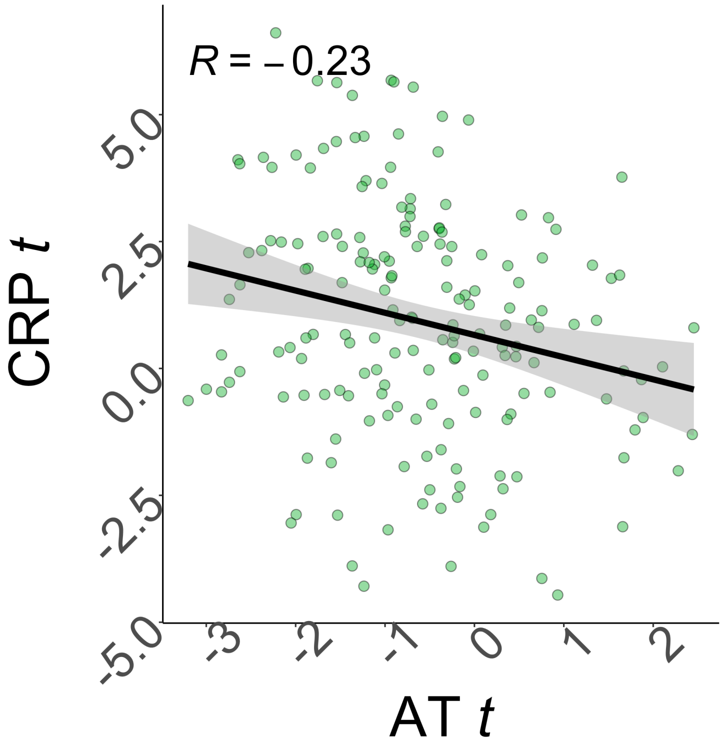

**Figure S9: Correlation of adult trauma and BMI effects on cortical thickness.** Scatterplot of effect ($t$-value) of CRP ($y$-axis) versus effect of AT ($x$-axis) on cortical thickness; each point represents one of 180 cortical areas. Spearman's correlation $\rho = -0.23$ over all areas, solid line is the regression of $t_{CRP}$ on $t_{AT}$

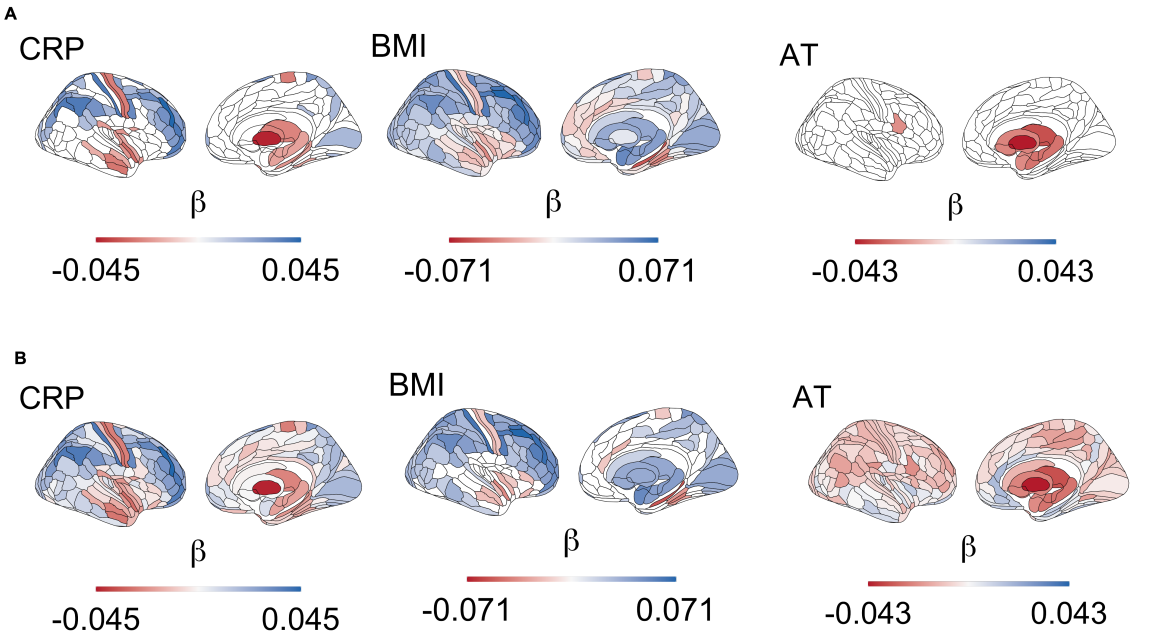

**Figure S10: Brain maps of independent linear relationships between adult trauma (AT), CRP or BMI and cortical thickness and subcortical volume.** **A** Each map shows the anatomical distribution of unthresholded linear regression $\beta$ coefficients. **B** $\beta$ maps thresholded for FDR<$0.05$ corrected significance.

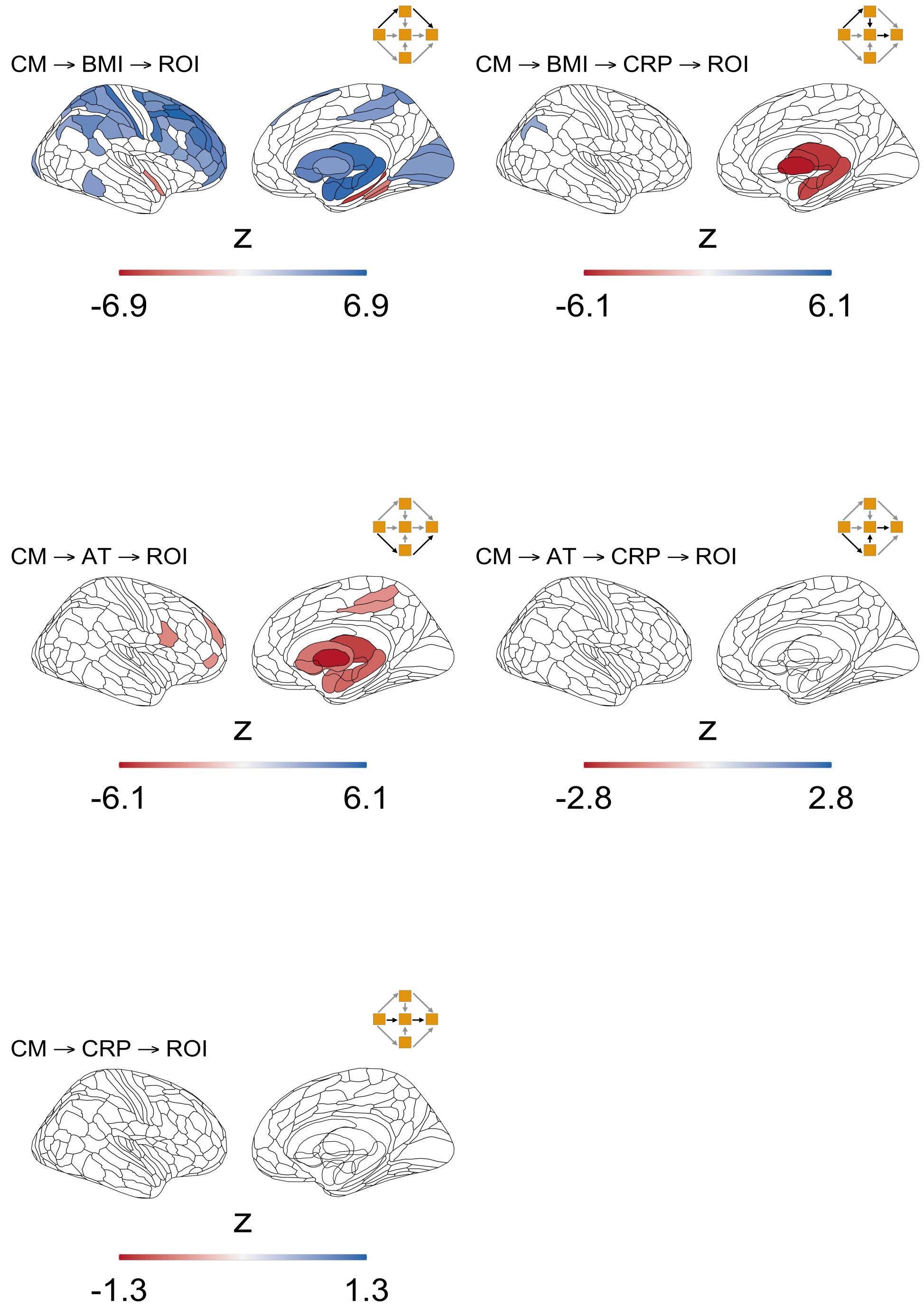

**Figure S11: Thresholded indirect ($z$) effects of childhood maltreatment on brain structure for all paths in the full model.**

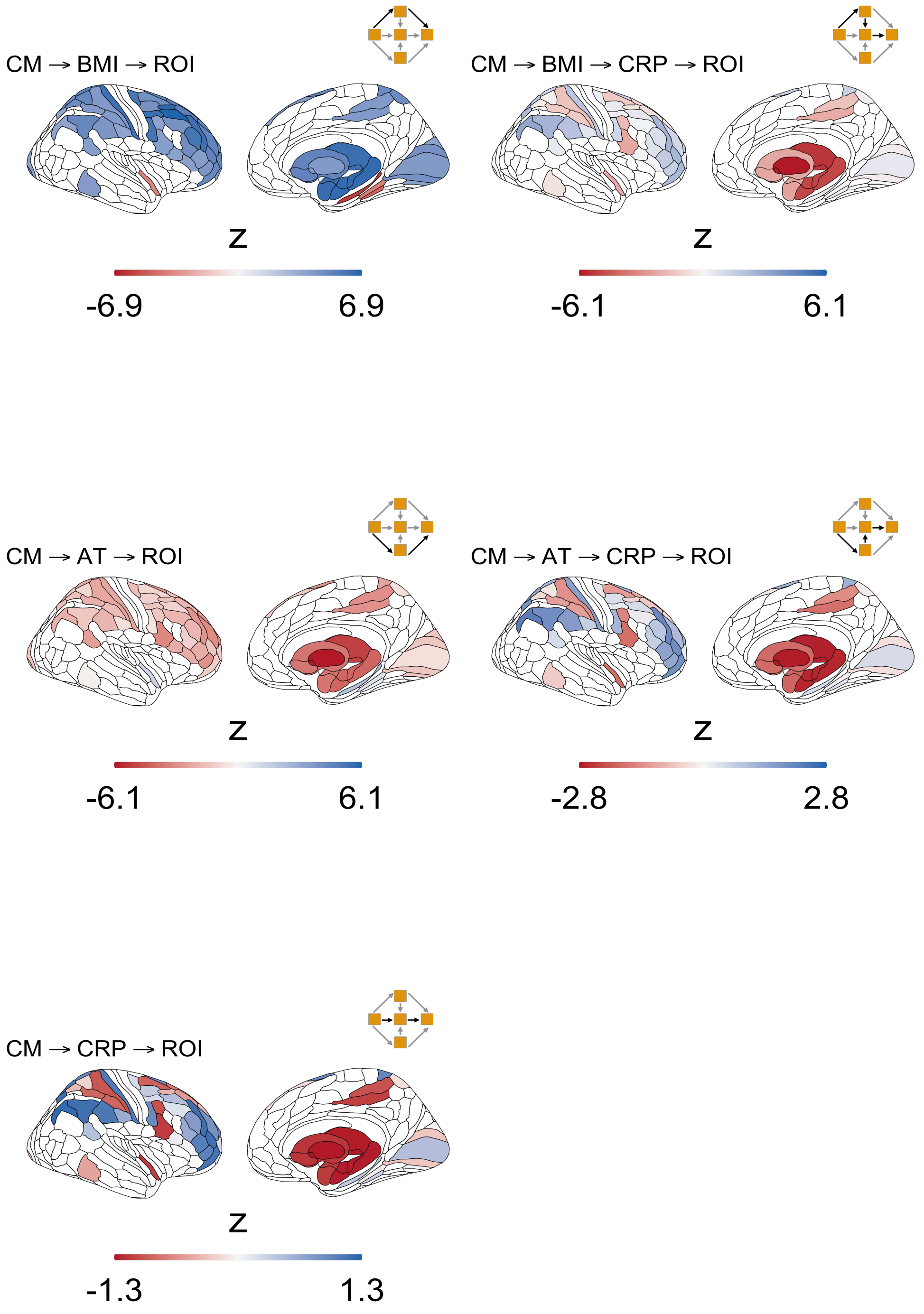

**Figure S12:** **Unthresholded indirect ($z$) effects of childhood maltreatment on brain structure for all paths in the full model**.

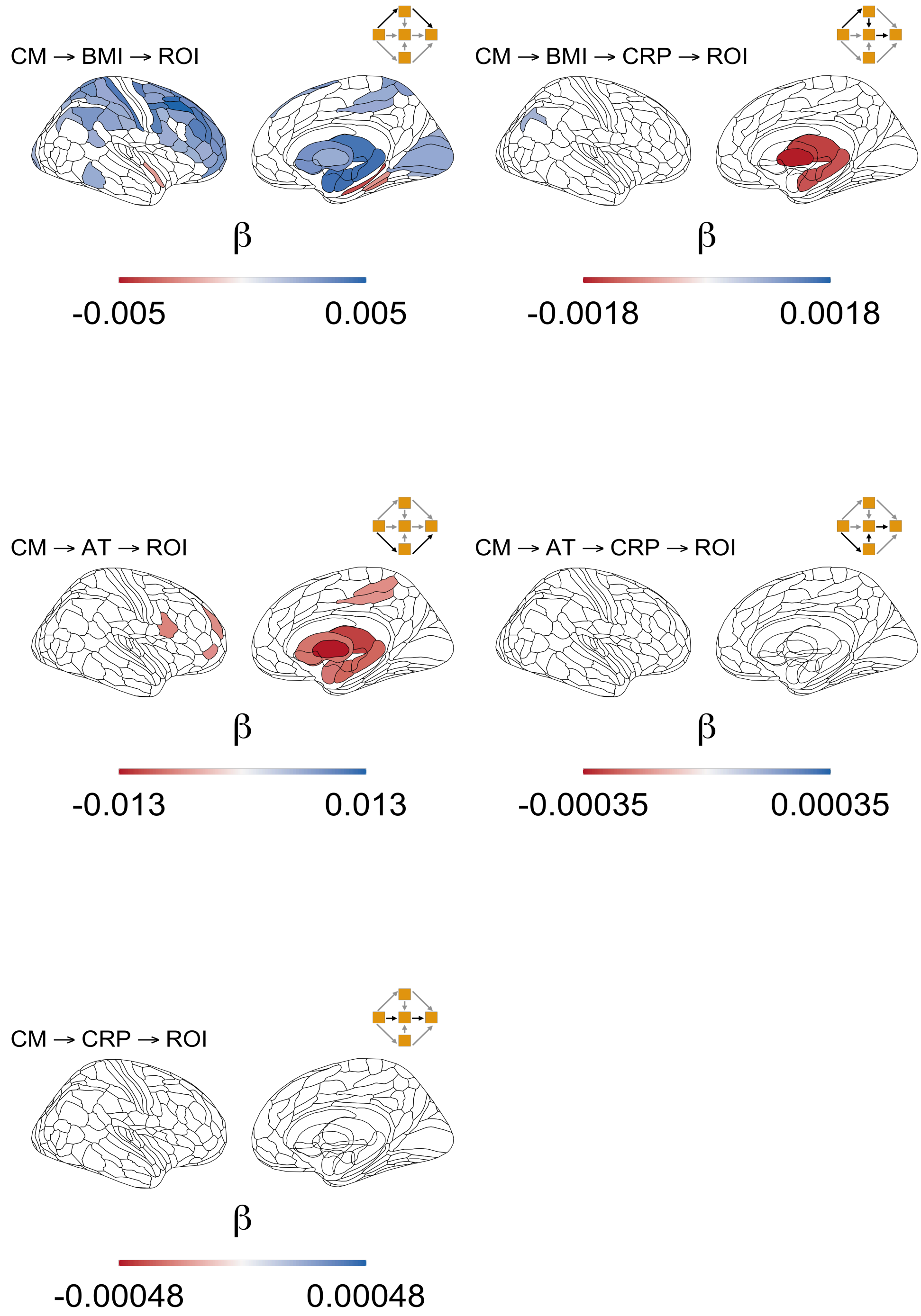

**Figure S13: Thresholded unstandardised indirect ($\beta$) effects of childhood maltreatment on brain structure for all paths in the full model.**

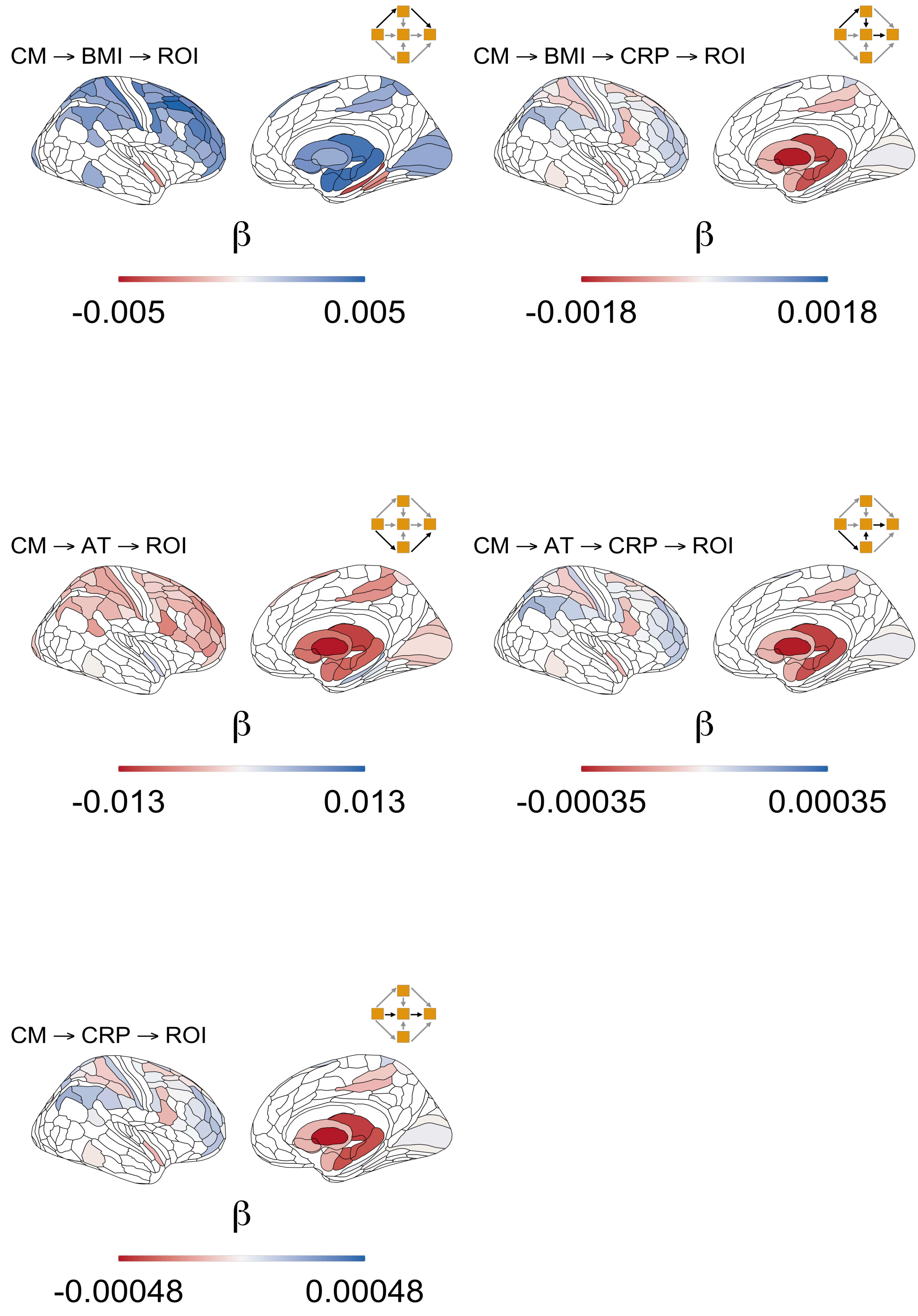

**Figure S14: Unthresholded unstandardised indirect ($\beta$) effects of childhood maltreatment on brain structure for all paths in the full model.**

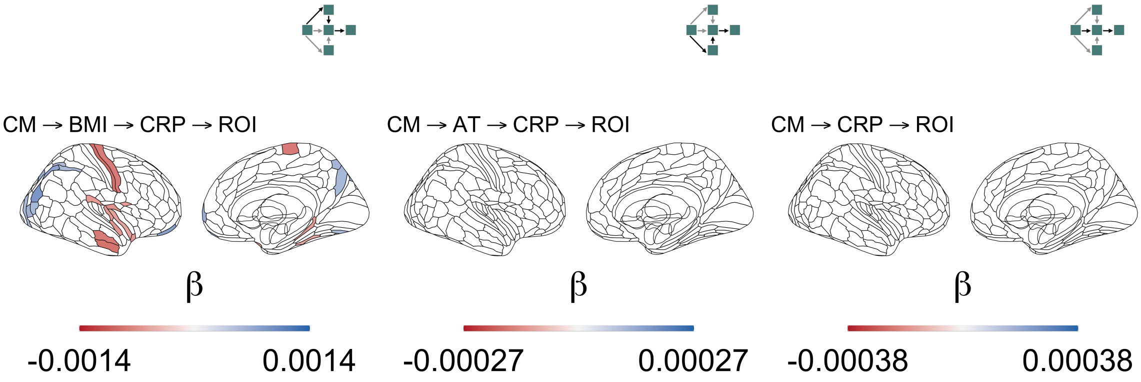

**Figure S15: Thresholded indirect ($z$) effects of childhood maltreatment on brain structure for all paths in the sparse model.**

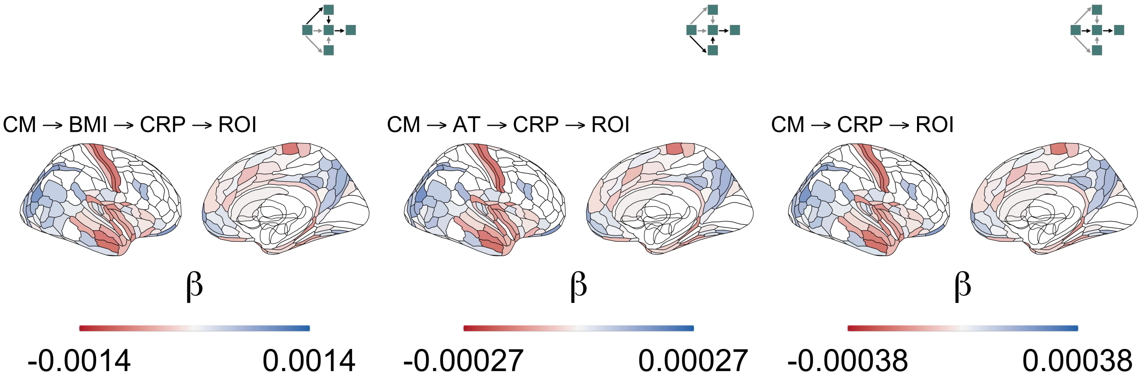

**Figure S16: Unthresholded indirect ($z$) effects of childhood maltreatment on brain structure for all paths in the sparse model.**

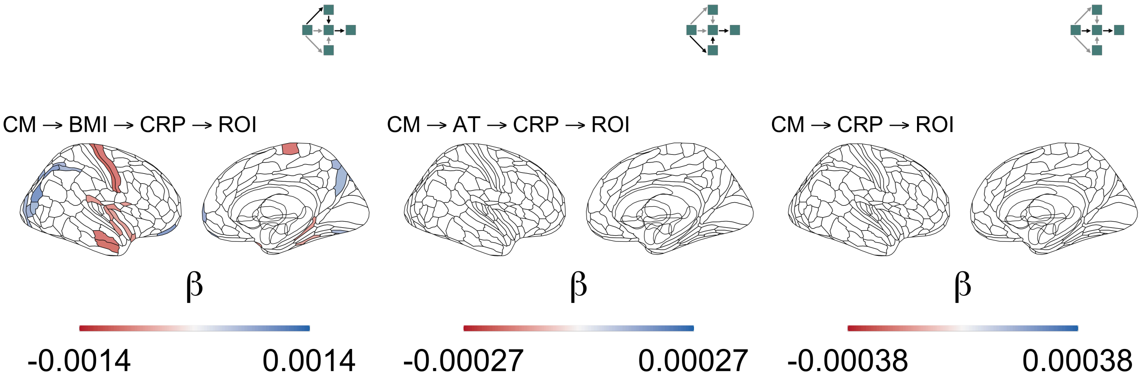

**Figure S17: Thresholded unstandardised indirect ($\beta$) effects of childhood maltreatment on brain structure for all paths in the sparse model.**

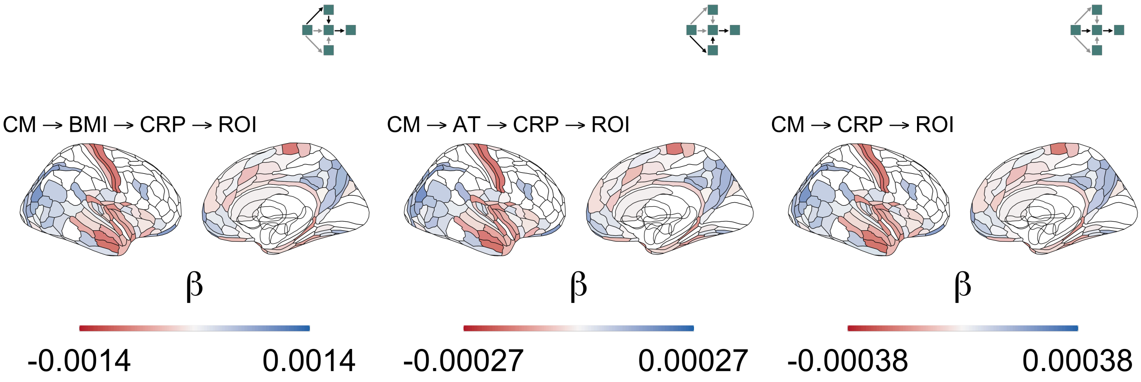

**Figure S18: Unthresholded unstandardised indirect ($\beta$) effects of childhood maltreatment on brain structure for all paths in the sparse model.**

### Supplementary Tables

| nuisance variable | $\beta$ | SE | t | $P-$value |
|---|---|---|---|---|
| **CRP** | | | | |
| deprivation | 0.015 | 0.003 | 5.801 | <0.0001 |
| age | 0.018 | 0.001 | 13.436 | <0.0001 |
| sex (male) | 0.236 | 0.104 | 2.270 | 0.0232 |
| age*sex (male) | -0.005 | 0.002 | -2.691 | 0.0071 |
| **BMI** | | | | |
| deprivation | 0.003 | 0.000 | 7.062 | <0.0001 |
| age | 0.001 | 0.000 | 7.588 | <0.0001 |
| sex (male) | 0.116 | 0.015 | 7.550 | <0.0001 |
| age*sex (male) | -0.001 | 0.000 | -4.808 | <0.0001 |
| **AT** | | | | |
| deprivation | 0.040 | 0.002 | 21.986 | <0.0001 |
| age | -0.001 | 0.001 | -0.775 | 0.4383 |
| sex (male) | -0.047 | 0.074 | -0.631 | 0.5279 |
| age*sex (male) | -0.003 | 0.001 | -1.867 | 0.0619 |
| **CM** | | | | |
| deprivation | 0.027 | 0.002 | 14.820 | <0.0001 |
| age | -0.001 | 0.001 | -1.182 | 0.2372 |
| sex (male) | 0.031 | 0.072 | 0.436 | 0.6629 |
| age*sex (male) | -0.001 | 0.001 | -0.846 | 0.3977 |

**Table ST1: Nuisance regression model results for immune, metabolic and psychosocial on UKB Imaging sample.**

| nuisance variable | $\beta$ | SE | t | $P-$value |
|---|---|---|---|---|
| **CRP** | | | | |
| deprivation | 0.013 | 0.001 | 12.390 | <0.0001 |
| age | 0.018 | 0.001 | 33.818 | <0.0001 |
| sex (male) | 0.181 | 0.045 | 4.051 | <0.0001 |
| age*sex (male) | -0.004 | 0.001 | -5.231 | <0.0001 |
| **BMI** | | | | |
| deprivation | 0.003 | 0.000 | 17.126 | <0.0001 |
| age | 0.001 | 0.000 | 17.751 | <0.0001 |
| sex (male) | 0.083 | 0.007 | 12.005 | <0.0001 |
| age*sex (male) | -0.001 | 0.000 | -6.235 | <0.0001 |
| **AT** | | | | |
| deprivation | 0.040 | 0.001 | 51.727 | 0.0000 |
| age | -0.003 | 0.000 | -7.290 | <0.0001 |
| sex (male) | -0.145 | 0.032 | -4.562 | <0.0001 |
| age*sex (male) | -0.001 | 0.001 | -0.920 | 0.3574 |
| **CM** | | | | |
| deprivation | 0.023 | 0.001 | 31.490 | <0.0001 |
| age | -0.003 | 0.000 | -8.481 | <0.0001 |
| sex (male) | -0.135 | 0.031 | -4.347 | <0.0001 |
| age*sex (male) | 0.002 | 0.001 | 3.763 | 0.0002 |

**Table ST2: Nuisance regression model results for immune, metabolic and psychosocial on the larger UKB sample.**

| Relationship | Predictor | Outcome | Path | $\beta$ | SE | Z | $P-$value | 95% CI |
|---|---|---|---|---|---|---|---|---|
| Direct | CM | BMI | a1 | 0.0721 | 0.0070 | 10.3433 | 0.0000 | 0.0584, 0.0857 |
| Direct | CM | CRP | a2 | 0.0084 | 0.0064 | 1.3130 | 0.1892 | -0.0042, 0.0210 |
| Direct | CM | AT | a3 | 0.3155 | 0.0065 | 48.8822 | 0.0000 | 0.3028, 0.3281 |
| Direct | BMI | CRP | b1 | 0.4340 | 0.0060 | 72.0471 | 0.0000 | 0.4222, 0.4458 |
| Direct | AT | CRP | b2 | 0.0193 | 0.0064 | 2.9991 | 0.0027 | 0.0067, 0.0319 |
| Indirect | CM | BMI$\rightarrow$CRP | a1*b1 | 0.0313 | 0.0030 | 10.2570 | 0.0000 | 0.0253, 0.0373 |
| Indirect | CM | AT$\rightarrow$CRP | a3*b2 | 0.0061 | 0.0020 | 2.9896 | 0.0028 | 0.0021, 0.0101 |

**Table ST3: Path model coefficients for the relationships between childhood maltreatment, adult trauma, BMI and CRP in the UKB MRI sample.**

| Relationship | Predictor | Outcome | Path | $\beta$ | SE | Z | $P-$value | 95% CI |
|---|---|---|---|---|---|---|---|---|
| Direct | CM | BMI | a1 | 0.0654 | 0.0030 | 21.5850 | 0.0000 | 0.0595, 0.0713 |
| Direct | CM | CRP | a2 | 0.0095 | 0.0028 | 3.4307 | 0.0006 | 0.0041, 0.0150 |
| Direct | CM | AT | a3 | 0.3289 | 0.0027 | 120.8475 | 0.0000 | 0.3235, 0.3342 |
| Direct | BMI | CRP | b1 | 0.4531 | 0.0026 | 176.8608 | 0.0000 | 0.4481, 0.4581 |
| Direct | AT | CRP | b2 | 0.0214 | 0.0028 | 7.7605 | $<$0.0001 | 0.0160, 0.0268 |
| Indirect | CM | BMI$\rightarrow$CRP | a1*b1 | 0.0296 | 0.0014 | 21.3995 | 0.0000 | 0.0269, 0.0323 |
| Indirect | CM | AT$\rightarrow$CRP | a3*b2 | 0.0070 | 0.0009 | 7.7408 | $<$0.0001 | 0.0053, 0.0088 |

**Table ST4: Path model coefficients for the relationships between childhood maltreatment, adult trauma, BMI and CRP in the larger UKB sample.**

**Table ST5:** **linear regression results at each brain area with CRP as the dependent variable and cortical thickness or subcortical volume as an independent variable**

| Brain area | layer | $\beta$ | SE | t-value | $P_{FDR}$ |
|---|---|---|---|---|---|
| Thalamus | subcortex | -0.0343446 | 0.0067791 | -5.0662711 | 0.0000361 |
| Caudate | subcortex | -0.0134896 | 0.0067838 | -1.9885112 | 0.2571494 |
| Putamen | subcortex | -0.0240733 | 0.0067820 | -3.5496031 | 0.0144598 |
| Pallidum | subcortex | -0.0427623 | 0.0067768 | -6.3101038 | 0.0000001 |
| Hippocampus | subcortex | -0.0275440 | 0.0067802 | -4.0623839 | 0.0022788 |
| Amygdala | subcortex | -0.0225451 | 0.0067811 | -3.3247045 | 0.0276320 |
| Accumbens | subcortex | -0.0338903 | 0.0067789 | -4.9993629 | 0.0000361 |
| V1 | cortex | -0.0026823 | 0.0067828 | -0.3954540 | 0.8712596 |
| MST | cortex | -0.0061870 | 0.0067827 | -0.9121820 | 0.6504152 |
| V6 | cortex | -0.0113643 | 0.0067824 | -1.6755612 | 0.3414714 |
| V2 | cortex | -0.0075707 | 0.0067826 | -1.1161897 | 0.5623793 |
| V3 | cortex | -0.0077400 | 0.0067826 | -1.1411486 | 0.5584055 |
| V4 | cortex | -0.0087003 | 0.0067826 | -1.2827432 | 0.4964930 |
| V8 | cortex | 0.0051557 | 0.0067827 | 0.7601246 | 0.7140984 |
| 4 | cortex | -0.0092737 | 0.0067825 | -1.3672875 | 0.4468037 |
| 3b | cortex | -0.0108675 | 0.0067846 | -1.6017949 | 0.3647018 |
| FEF | cortex | -0.0052475 | 0.0067827 | -0.7736608 | 0.7140984 |
| PEF | cortex | -0.0083379 | 0.0067826 | -1.2293058 | 0.5055245 |
| 55b | cortex | -0.0063611 | 0.0067827 | -0.9378386 | 0.6504152 |
| V3A | cortex | -0.0003910 | 0.0067828 | -0.0576433 | 0.9695882 |
| RSC | cortex | -0.0019627 | 0.0067832 | -0.2893516 | 0.8829476 |
| POS2 | cortex | -0.0052331 | 0.0067827 | -0.7715347 | 0.7140984 |
| V7 | cortex | -0.0068088 | 0.0067827 | -1.0038553 | 0.6275631 |
| IPS1 | cortex | -0.0021771 | 0.0067828 | -0.3209802 | 0.8743274 |
| FFC | cortex | 0.0021131 | 0.0067828 | 0.3115376 | 0.8743274 |
| V3B | cortex | -0.0020956 | 0.0067828 | -0.3089526 | 0.8743274 |
| LO1 | cortex | 0.0056191 | 0.0067827 | 0.8284472 | 0.6863850 |
| LO2 | cortex | 0.0045209 | 0.0067827 | 0.6665328 | 0.7436970 |
| PIT | cortex | -0.0102186 | 0.0067825 | -1.5066175 | 0.3915817 |
| MT | cortex | -0.0015185 | 0.0067828 | -0.2238754 | 0.9051420 |
| A1 | cortex | 0.0092637 | 0.0067825 | 1.3658161 | 0.4468037 |
| PSL | cortex | -0.0134039 | 0.0067822 | -1.9763341 | 0.2571494 |
| SFL | cortex | -0.0064590 | 0.0067827 | -0.9522753 | 0.6504152 |
| PCV | cortex | -0.0100389 | 0.0067825 | -1.4801164 | 0.3954582 |
| STV | cortex | -0.0154606 | 0.0067820 | -2.2796464 | 0.1833266 |
| 7Pm | cortex | 0.0061865 | 0.0067827 | 0.9120961 | 0.6504152 |
| 7m | cortex | 0.0089561 | 0.0067825 | 1.3204583 | 0.4717857 |
| POS1 | cortex | -0.0063129 | 0.0067827 | -0.9307364 | 0.6504152 |
| 23d | cortex | -0.0075756 | 0.0067841 | -1.1166775 | 0.5623793 |
| v23ab | cortex | 0.0006429 | 0.0067828 | 0.0947776 | 0.9604448 |
| d23ab | cortex | -0.0043473 | 0.0067831 | -0.6408993 | 0.7544673 |
| 31pv | cortex | -0.0068322 | 0.0067827 | -1.0073058 | 0.6275631 |
| 5m | cortex | -0.0025308 | 0.0067828 | -0.3731237 | 0.8712596 |
| 5mv | cortex | -0.0191878 | 0.0067816 | -2.8294042 | 0.0873442 |
| 23c | cortex | -0.0191864 | 0.0067816 | -2.8291941 | 0.0873442 |

**Table ST5:** **linear regression results at each brain area with CRP as the dependent variable and cortical thickness or subcortical volume as an independent variable** *(continued)*

| Brain area | layer | $\beta$ | SE | t-value | $P_{FDR}$ |
|---|---|---|---|---|---|
| 5L | cortex | -0.0135293 | 0.0067822 | -1.9948183 | 0.2571494 |
| 24dd | cortex | -0.0177978 | 0.0067817 | -2.6243752 | 0.1018327 |
| 24dv | cortex | -0.0095565 | 0.0067825 | -1.4089887 | 0.4368452 |
| 7AL | cortex | -0.0114492 | 0.0067824 | -1.6880866 | 0.3414714 |
| SCEF | cortex | -0.0046428 | 0.0067827 | -0.6845000 | 0.7326642 |
| 6ma | cortex | -0.0160117 | 0.0067819 | -2.3609321 | 0.1624036 |
| 7Am | cortex | -0.0026092 | 0.0067828 | -0.3846798 | 0.8712596 |
| 7PL | cortex | -0.0092632 | 0.0067825 | -1.3657506 | 0.4468037 |
| 7PC | cortex | -0.0038723 | 0.0067828 | -0.5709017 | 0.8047689 |
| LIPv | cortex | -0.0084362 | 0.0067826 | -1.2438057 | 0.5055245 |
| VIP | cortex | -0.0057625 | 0.0067827 | -0.8495840 | 0.6820270 |
| MIP | cortex | -0.0083808 | 0.0067826 | -1.2356382 | 0.5055245 |
| 1 | cortex | -0.0119166 | 0.0067823 | -1.7570086 | 0.3140423 |
| 2 | cortex | -0.0139923 | 0.0067822 | -2.0631020 | 0.2431033 |
| 3a | cortex | -0.0083857 | 0.0067826 | -1.2363539 | 0.5055245 |
| 6d | cortex | -0.0082262 | 0.0067826 | -1.2128409 | 0.5135746 |
| 6mp | cortex | -0.0129269 | 0.0067823 | -1.9059887 | 0.2726984 |
| 6v | cortex | -0.0004454 | 0.0067828 | -0.0656723 | 0.9695882 |
| p24pr | cortex | -0.0032370 | 0.0067828 | -0.4772356 | 0.8468744 |
| 33pr | cortex | 0.0100207 | 0.0067825 | 1.4774352 | 0.3954582 |
| a24pr | cortex | 0.0057384 | 0.0067827 | 0.8460303 | 0.6820270 |
| p32pr | cortex | -0.0036090 | 0.0067830 | -0.5320661 | 0.8289377 |
| a24 | cortex | 0.0166490 | 0.0067820 | 2.4548827 | 0.1456455 |
| d32 | cortex | -0.0068137 | 0.0067827 | -1.0045732 | 0.6275631 |
| 8BM | cortex | -0.0034552 | 0.0067828 | -0.5094139 | 0.8393925 |
| p32 | cortex | -0.0016495 | 0.0067828 | -0.2431885 | 0.9014136 |
| 10r | cortex | 0.0033628 | 0.0067832 | 0.4957591 | 0.8402390 |
| 47m | cortex | -0.0008294 | 0.0067828 | -0.1222725 | 0.9536834 |
| 8Av | cortex | -0.0024311 | 0.0067828 | -0.3584145 | 0.8712596 |
| 8Ad | cortex | -0.0100225 | 0.0067825 | -1.4777031 | 0.3954582 |
| 9m | cortex | -0.0058499 | 0.0067827 | -0.8624668 | 0.6788628 |
| 8BL | cortex | -0.0062599 | 0.0067827 | -0.9229206 | 0.6504152 |
| 9p | cortex | -0.0171253 | 0.0067818 | -2.5251813 | 0.1272800 |
| 10d | cortex | -0.0048741 | 0.0067828 | -0.7186020 | 0.7277287 |
| 8C | cortex | -0.0104541 | 0.0067824 | -1.5413506 | 0.3794707 |
| 44 | cortex | -0.0126243 | 0.0067823 | -1.8613656 | 0.2726984 |
| 45 | cortex | -0.0128380 | 0.0067823 | -1.8928741 | 0.2726984 |
| 47l | cortex | 0.0023660 | 0.0067828 | 0.3488218 | 0.8712596 |
| a47r | cortex | -0.0046431 | 0.0067827 | -0.6845403 | 0.7326642 |
| 6r | cortex | -0.0217190 | 0.0067812 | -3.2028126 | 0.0364085 |
| IFJa | cortex | -0.0017187 | 0.0067828 | -0.2533946 | 0.9011662 |
| IFJp | cortex | 0.0025317 | 0.0067828 | 0.3732496 | 0.8712596 |
| IFSp | cortex | -0.0048479 | 0.0067827 | -0.7147441 | 0.7277287 |
| IFSa | cortex | -0.0124491 | 0.0067823 | -1.8355367 | 0.2823694 |
| p9-46v | cortex | -0.0090460 | 0.0067825 | -1.3337129 | 0.4670183 |

**Table ST5:** **linear regression results at each brain area with CRP as the dependent variable and cortical thickness or subcortical volume as an independent variable** *(continued)*

| Brain area | layer | $\beta$ | SE | t-value | $P_{FDR}$ |
|---|---|---|---|---|---|
| 46 | cortex | -0.0150781 | 0.0067820 | -2.2232313 | 0.1960541 |
| a9-46v | cortex | -0.0063168 | 0.0067827 | -0.9313165 | 0.6504152 |
| 9-46d | cortex | -0.0179443 | 0.0067817 | -2.6459791 | 0.1018327 |
| 9a | cortex | -0.0153606 | 0.0067820 | -2.2649011 | 0.1833266 |
| 10v | cortex | 0.0026768 | 0.0067828 | 0.3946474 | 0.8712596 |
| a10p | cortex | 0.0005453 | 0.0067830 | 0.0803908 | 0.9669524 |
| 10pp | cortex | 0.0035723 | 0.0067828 | 0.5266659 | 0.8289377 |
| 11l | cortex | 0.0111894 | 0.0067824 | 1.6497671 | 0.3428508 |
| 13l | cortex | 0.0103882 | 0.0067825 | 1.5316300 | 0.3794707 |
| OFC | cortex | 0.0113444 | 0.0067826 | 1.6725642 | 0.3414714 |
| 47s | cortex | -0.0013689 | 0.0067828 | -0.2018194 | 0.9133209 |
| LIPd | cortex | -0.0146524 | 0.0067821 | -2.1604554 | 0.2129606 |
| 6a | cortex | -0.0046994 | 0.0067827 | -0.6928469 | 0.7326642 |
| i6-8 | cortex | -0.0070213 | 0.0067827 | -1.0351917 | 0.6182092 |
| s6-8 | cortex | -0.0086581 | 0.0067826 | -1.2765260 | 0.4964930 |
| 43 | cortex | -0.0074222 | 0.0067826 | -1.0942907 | 0.5753709 |
| OP4 | cortex | 0.0043168 | 0.0067828 | 0.6364406 | 0.7544673 |
| OP1 | cortex | -0.0015381 | 0.0067828 | -0.2267703 | 0.9051420 |
| OP2-3 | cortex | -0.0105290 | 0.0067824 | -1.5523959 | 0.3794707 |
| 52 | cortex | -0.0103839 | 0.0067830 | -1.5308791 | 0.3794707 |
| RI | cortex | -0.0135249 | 0.0067822 | -1.9941751 | 0.2571494 |
| PFcm | cortex | -0.0203349 | 0.0067814 | -2.9986249 | 0.0634650 |
| PoI2 | cortex | 0.0019446 | 0.0067828 | 0.2866935 | 0.8829476 |
| TA2 | cortex | 0.0007071 | 0.0067828 | 0.1042461 | 0.9579825 |
| FOP4 | cortex | -0.0144965 | 0.0067821 | -2.1374677 | 0.2175284 |
| MI | cortex | 0.0126769 | 0.0067823 | 1.8691247 | 0.2726984 |
| Pir | cortex | -0.0139087 | 0.0067822 | -2.0507778 | 0.2431033 |
| AVI | cortex | 0.0037609 | 0.0067828 | 0.5544765 | 0.8144461 |
| AAIC | cortex | -0.0024006 | 0.0067828 | -0.3539308 | 0.8712596 |
| FOP1 | cortex | -0.0025410 | 0.0067828 | -0.3746246 | 0.8712596 |
| FOP3 | cortex | -0.0056662 | 0.0067831 | -0.8353324 | 0.6860176 |
| FOP2 | cortex | -0.0065657 | 0.0067827 | -0.9680166 | 0.6501731 |
| PFt | cortex | -0.0097912 | 0.0067825 | -1.4435925 | 0.4154974 |
| AIP | cortex | -0.0122334 | 0.0067823 | -1.8037233 | 0.2943295 |
| EC | cortex | 0.0121856 | 0.0067823 | 1.7966700 | 0.2943295 |
| PreS | cortex | 0.0012400 | 0.0067828 | 0.1828174 | 0.9188180 |
| H | cortex | 0.0112494 | 0.0067824 | 1.6586141 | 0.3428508 |
| ProS | cortex | -0.0131159 | 0.0067822 | -1.9338654 | 0.2726984 |
| PeEc | cortex | 0.0027639 | 0.0067828 | 0.4074823 | 0.8712596 |
| STGa | cortex | -0.0126748 | 0.0067823 | -1.8688190 | 0.2726984 |
| PBelt | cortex | -0.0079694 | 0.0067826 | -1.1749776 | 0.5343226 |
| A5 | cortex | -0.0070177 | 0.0067827 | -1.0346589 | 0.6182092 |
| PHA1 | cortex | 0.0032222 | 0.0067828 | 0.4750504 | 0.8468744 |
| PHA3 | cortex | -0.0012818 | 0.0067828 | -0.1889738 | 0.9188180 |
| STSda | cortex | -0.0033831 | 0.0067828 | -0.4987745 | 0.8402390 |

**Table ST5:** **linear regression results at each brain area with CRP as the dependent variable and cortical thickness or subcortical volume as an independent variable** *(continued)*

| Brain area | layer | $\beta$ | SE | t-value | $P_{FDR}$ |
|---|---|---|---|---|---|
| STSdp | cortex | 0.0000911 | 0.0067828 | 0.0134254 | 0.9957617 |
| STSvp | cortex | 0.0021693 | 0.0067828 | 0.3198179 | 0.8743274 |
| TGd | cortex | -0.0053294 | 0.0067827 | -0.7857381 | 0.7140984 |
| TE1a | cortex | 0.0051170 | 0.0067827 | 0.7544161 | 0.7140984 |
| TE1p | cortex | 0.0000342 | 0.0067828 | 0.0050428 | 0.9959765 |
| TE2a | cortex | 0.0075660 | 0.0067826 | 1.1154988 | 0.5623793 |
| TF | cortex | 0.0154650 | 0.0067820 | 2.2802949 | 0.1833266 |
| TE2p | cortex | 0.0031807 | 0.0067828 | 0.4689323 | 0.8468744 |
| PHT | cortex | 0.0051210 | 0.0067827 | 0.7550067 | 0.7140984 |
| PH | cortex | -0.0127840 | 0.0067823 | -1.8849104 | 0.2726984 |
| TPOJ1 | cortex | -0.0094653 | 0.0067825 | -1.3955461 | 0.4413890 |
| TPOJ2 | cortex | -0.0185936 | 0.0067816 | -2.7417607 | 0.0955915 |
| TPOJ3 | cortex | -0.0079845 | 0.0067826 | -1.1771969 | 0.5343226 |
| DVT | cortex | -0.0161369 | 0.0067819 | -2.3793984 | 0.1622178 |
| PGp | cortex | -0.0027552 | 0.0067828 | -0.4062013 | 0.8712596 |
| IP2 | cortex | -0.0114892 | 0.0067824 | -1.6939758 | 0.3414714 |
| IP1 | cortex | -0.0085231 | 0.0067826 | -1.2566215 | 0.5055245 |
| IP0 | cortex | -0.0047627 | 0.0067827 | -0.7021832 | 0.7326642 |
| PFop | cortex | -0.0026610 | 0.0067828 | -0.3923232 | 0.8712596 |
| PF | cortex | -0.0104788 | 0.0067824 | -1.5449946 | 0.3794707 |
| PFm | cortex | -0.0061107 | 0.0067827 | -0.9009310 | 0.6547404 |
| PGi | cortex | -0.0177909 | 0.0067817 | -2.6233539 | 0.1018327 |
| PGs | cortex | -0.0104427 | 0.0067824 | -1.5396581 | 0.3794707 |
| V6A | cortex | -0.0048869 | 0.0067827 | -0.7204846 | 0.7277287 |
| VMV1 | cortex | -0.0185871 | 0.0067816 | -2.7407879 | 0.0955915 |
| VMV3 | cortex | 0.0048682 | 0.0067827 | 0.7177313 | 0.7277287 |
| PHA2 | cortex | -0.0011028 | 0.0067828 | -0.1625820 | 0.9252772 |
| V4t | cortex | -0.0000813 | 0.0067828 | -0.0119862 | 0.9957617 |
| FST | cortex | -0.0020949 | 0.0067828 | -0.3088499 | 0.8743274 |
| V3CD | cortex | -0.0007069 | 0.0067828 | -0.1042155 | 0.9579825 |
| LO3 | cortex | -0.0024573 | 0.0067828 | -0.3622796 | 0.8712596 |
| VMV2 | cortex | 0.0003991 | 0.0067828 | 0.0588364 | 0.9695882 |
| 31pd | cortex | -0.0148484 | 0.0067821 | -2.1893561 | 0.2055674 |
| 31a | cortex | -0.0011554 | 0.0067828 | -0.1703478 | 0.9240346 |
| VVC | cortex | 0.0127968 | 0.0067823 | 1.8868038 | 0.2726984 |
| 25 | cortex | 0.0113263 | 0.0067826 | 1.6698946 | 0.3414714 |
| s32 | cortex | 0.0142732 | 0.0067821 | 2.1045260 | 0.2279081 |
| pOFC | cortex | 0.0031432 | 0.0067828 | 0.4634017 | 0.8468744 |
| PoI1 | cortex | -0.0039267 | 0.0067828 | -0.5789183 | 0.8031725 |
| Ig | cortex | -0.0065558 | 0.0067827 | -0.9665522 | 0.6501731 |
| FOP5 | cortex | -0.0044274 | 0.0067828 | -0.6527479 | 0.7508134 |
| p10p | cortex | -0.0055113 | 0.0067827 | -0.8125488 | 0.6953824 |
| p47r | cortex | -0.0178048 | 0.0067817 | -2.6253955 | 0.1018327 |
| TGv | cortex | 0.0023276 | 0.0067828 | 0.3431547 | 0.8712596 |
| MBelt | cortex | 0.0024815 | 0.0067828 | 0.3658529 | 0.8712596 |

**Table ST5:** **linear regression results at each brain area with CRP as the dependent variable and cortical thickness or subcortical volume as an independent variable** *(continued)*

| Brain area | layer | $\beta$ | SE | t-value | $P_{FDR}$ |
|---|---|---|---|---|---|
| LBelt | cortex | -0.0016323 | 0.0067828 | -0.2406532 | 0.9014136 |
| A4 | cortex | -0.0060323 | 0.0067827 | -0.8893734 | 0.6594612 |
| STSva | cortex | -0.0017739 | 0.0067828 | -0.2615279 | 0.8995130 |
| TE1m | cortex | 0.0110191 | 0.0067824 | 1.6246650 | 0.3544448 |
| PI | cortex | 0.0063200 | 0.0067827 | 0.9317801 | 0.6504152 |
| a32pr | cortex | -0.0014223 | 0.0067828 | -0.2096861 | 0.9119418 |
| p24 | cortex | 0.0165307 | 0.0067819 | 2.4374747 | 0.1456455 |

**Table ST6:** **linear regression results at each brain area with BMI as the dependent variable and cortical thickness or subcortical volume as an independent variable**

| Brain area | layer | $\beta$ | SE | t-value | $P_{FDR}$ |
|---|---|---|---|---|---|
| Thalamus | subcortex | -0.0343446 | 0.0067791 | -5.0662711 | 0.0000361 |
| Caudate | subcortex | -0.0134896 | 0.0067838 | -1.9885112 | 0.2571494 |
| Putamen | subcortex | -0.0240733 | 0.0067820 | -3.5496031 | 0.0144598 |
| Pallidum | subcortex | -0.0427623 | 0.0067768 | -6.3101038 | 0.0000001 |
| Hippocampus | subcortex | -0.0275440 | 0.0067802 | -4.0623839 | 0.0022788 |
| Amygdala | subcortex | -0.0225451 | 0.0067811 | -3.3247045 | 0.0276320 |
| Accumbens | subcortex | -0.0338903 | 0.0067789 | -4.9993629 | 0.0000361 |
| V1 | cortex | -0.0026823 | 0.0067828 | -0.3954540 | 0.8712596 |
| MST | cortex | -0.0061870 | 0.0067827 | -0.9121820 | 0.6504152 |
| V6 | cortex | -0.0113643 | 0.0067824 | -1.6755612 | 0.3414714 |
| V2 | cortex | -0.0075707 | 0.0067826 | -1.1161897 | 0.5623793 |
| V3 | cortex | -0.0077400 | 0.0067826 | -1.1411486 | 0.5584055 |
| V4 | cortex | -0.0087003 | 0.0067826 | -1.2827432 | 0.4964930 |
| V8 | cortex | 0.0051557 | 0.0067827 | 0.7601246 | 0.7140984 |
| 4 | cortex | -0.0092737 | 0.0067825 | -1.3672875 | 0.4468037 |
| 3b | cortex | -0.0108675 | 0.0067846 | -1.6017949 | 0.3647018 |
| FEF | cortex | -0.0052475 | 0.0067827 | -0.7736608 | 0.7140984 |
| PEF | cortex | -0.0083379 | 0.0067826 | -1.2293058 | 0.5055245 |
| 55b | cortex | -0.0063611 | 0.0067827 | -0.9378386 | 0.6504152 |
| V3A | cortex | -0.0003910 | 0.0067828 | -0.0576433 | 0.9695882 |
| RSC | cortex | -0.0019627 | 0.0067832 | -0.2893516 | 0.8829476 |
| POS2 | cortex | -0.0052331 | 0.0067827 | -0.7715347 | 0.7140984 |
| V7 | cortex | -0.0068088 | 0.0067827 | -1.0038553 | 0.6275631 |
| IPS1 | cortex | -0.0021771 | 0.0067828 | -0.3209802 | 0.8743274 |
| FFC | cortex | 0.0021131 | 0.0067828 | 0.3115376 | 0.8743274 |
| V3B | cortex | -0.0020956 | 0.0067828 | -0.3089526 | 0.8743274 |
| LO1 | cortex | 0.0056191 | 0.0067827 | 0.8284472 | 0.6863850 |
| LO2 | cortex | 0.0045209 | 0.0067827 | 0.6665328 | 0.7436970 |
| PIT | cortex | -0.0102186 | 0.0067825 | -1.5066175 | 0.3915817 |
| MT | cortex | -0.0015185 | 0.0067828 | -0.2238754 | 0.9051420 |
| A1 | cortex | 0.0092637 | 0.0067825 | 1.3658161 | 0.4468037 |
| PSL | cortex | -0.0134039 | 0.0067822 | -1.9763341 | 0.2571494 |
| SFL | cortex | -0.0064590 | 0.0067827 | -0.9522753 | 0.6504152 |
| PCV | cortex | -0.0100389 | 0.0067825 | -1.4801164 | 0.3954582 |
| STV | cortex | -0.0154606 | 0.0067820 | -2.2796464 | 0.1833266 |
| 7Pm | cortex | 0.0061865 | 0.0067827 | 0.9120961 | 0.6504152 |
| 7m | cortex | 0.0089561 | 0.0067825 | 1.3204583 | 0.4717857 |
| POS1 | cortex | -0.0063129 | 0.0067827 | -0.9307364 | 0.6504152 |
| 23d | cortex | -0.0075756 | 0.0067841 | -1.1166775 | 0.5623793 |
| v23ab | cortex | 0.0006429 | 0.0067828 | 0.0947776 | 0.9604448 |
| d23ab | cortex | -0.0043473 | 0.0067831 | -0.6408993 | 0.7544673 |
| 31pv | cortex | -0.0068322 | 0.0067827 | -1.0073058 | 0.6275631 |
| 5m | cortex | -0.0025308 | 0.0067828 | -0.3731237 | 0.8712596 |
| 5mv | cortex | -0.0191878 | 0.0067816 | -2.8294042 | 0.0873442 |
| 23c | cortex | -0.0191864 | 0.0067816 | -2.8291941 | 0.0873442 |

**Table ST6:** **linear regression results at each brain area with BMI as the dependent variable and cortical thickness or subcortical volume as an independent variable** *(continued)*

| Brain area | layer | $\beta$ | SE | t-value | $P_{FDR}$ |
|---|---|---|---|---|---|
| 5L | cortex | -0.0135293 | 0.0067822 | -1.9948183 | 0.2571494 |
| 24dd | cortex | -0.0177978 | 0.0067817 | -2.6243752 | 0.1018327 |
| 24dv | cortex | -0.0095565 | 0.0067825 | -1.4089887 | 0.4368452 |
| 7AL | cortex | -0.0114492 | 0.0067824 | -1.6880866 | 0.3414714 |
| SCEF | cortex | -0.0046428 | 0.0067827 | -0.6845000 | 0.7326642 |
| 6ma | cortex | -0.0160117 | 0.0067819 | -2.3609321 | 0.1624036 |
| 7Am | cortex | -0.0026092 | 0.0067828 | -0.3846798 | 0.8712596 |
| 7PL | cortex | -0.0092632 | 0.0067825 | -1.3657506 | 0.4468037 |
| 7PC | cortex | -0.0038723 | 0.0067828 | -0.5709017 | 0.8047689 |
| LIPv | cortex | -0.0084362 | 0.0067826 | -1.2438057 | 0.5055245 |
| VIP | cortex | -0.0057625 | 0.0067827 | -0.8495840 | 0.6820270 |
| MIP | cortex | -0.0083808 | 0.0067826 | -1.2356382 | 0.5055245 |
| 1 | cortex | -0.0119166 | 0.0067823 | -1.7570086 | 0.3140423 |
| 2 | cortex | -0.0139923 | 0.0067822 | -2.0631020 | 0.2431033 |
| 3a | cortex | -0.0083857 | 0.0067826 | -1.2363539 | 0.5055245 |
| 6d | cortex | -0.0082262 | 0.0067826 | -1.2128409 | 0.5135746 |
| 6mp | cortex | -0.0129269 | 0.0067823 | -1.9059887 | 0.2726984 |
| 6v | cortex | -0.0004454 | 0.0067828 | -0.0656723 | 0.9695882 |
| p24pr | cortex | -0.0032370 | 0.0067828 | -0.4772356 | 0.8468744 |
| 33pr | cortex | 0.0100207 | 0.0067825 | 1.4774352 | 0.3954582 |
| a24pr | cortex | 0.0057384 | 0.0067827 | 0.8460303 | 0.6820270 |
| p32pr | cortex | -0.0036090 | 0.0067830 | -0.5320661 | 0.8289377 |
| a24 | cortex | 0.0166490 | 0.0067820 | 2.4548827 | 0.1456455 |
| d32 | cortex | -0.0068137 | 0.0067827 | -1.0045732 | 0.6275631 |
| 8BM | cortex | -0.0034552 | 0.0067828 | -0.5094139 | 0.8393925 |
| p32 | cortex | -0.0016495 | 0.0067828 | -0.2431885 | 0.9014136 |
| 10r | cortex | 0.0033628 | 0.0067832 | 0.4957591 | 0.8402390 |
| 47m | cortex | -0.0008294 | 0.0067828 | -0.1222725 | 0.9536834 |
| 8Av | cortex | -0.0024311 | 0.0067828 | -0.3584145 | 0.8712596 |
| 8Ad | cortex | -0.0100225 | 0.0067825 | -1.4777031 | 0.3954582 |
| 9m | cortex | -0.0058499 | 0.0067827 | -0.8624668 | 0.6788628 |
| 8BL | cortex | -0.0062599 | 0.0067827 | -0.9229206 | 0.6504152 |
| 9p | cortex | -0.0171253 | 0.0067818 | -2.5251813 | 0.1272800 |
| 10d | cortex | -0.0048741 | 0.0067828 | -0.7186020 | 0.7277287 |
| 8C | cortex | -0.0104541 | 0.0067824 | -1.5413506 | 0.3794707 |
| 44 | cortex | -0.0126243 | 0.0067823 | -1.8613656 | 0.2726984 |
| 45 | cortex | -0.0128380 | 0.0067823 | -1.8928741 | 0.2726984 |
| 47l | cortex | 0.0023660 | 0.0067828 | 0.3488218 | 0.8712596 |
| a47r | cortex | -0.0046431 | 0.0067827 | -0.6845403 | 0.7326642 |
| 6r | cortex | -0.0217190 | 0.0067812 | -3.2028126 | 0.0364085 |
| IFJa | cortex | -0.0017187 | 0.0067828 | -0.2533946 | 0.9011662 |
| IFJp | cortex | 0.0025317 | 0.0067828 | 0.3732496 | 0.8712596 |
| IFSp | cortex | -0.0048479 | 0.0067827 | -0.7147441 | 0.7277287 |
| IFSa | cortex | -0.0124491 | 0.0067823 | -1.8355367 | 0.2823694 |
| p9-46v | cortex | -0.0090460 | 0.0067825 | -1.3337129 | 0.4670183 |

**Table ST6:** **linear regression results at each brain area with BMI as the dependent variable and cortical thickness or subcortical volume as an independent variable** *(continued)*

| Brain area | layer | $\beta$ | SE | t-value | $P_{FDR}$ |
|---|---|---|---|---|---|
| 46 | cortex | -0.0150781 | 0.0067820 | -2.2232313 | 0.1960541 |
| a9-46v | cortex | -0.0063168 | 0.0067827 | -0.9313165 | 0.6504152 |
| 9-46d | cortex | -0.0179443 | 0.0067817 | -2.6459791 | 0.1018327 |
| 9a | cortex | -0.0153606 | 0.0067820 | -2.2649011 | 0.1833266 |
| 10v | cortex | 0.0026768 | 0.0067828 | 0.3946474 | 0.8712596 |
| a10p | cortex | 0.0005453 | 0.0067830 | 0.0803908 | 0.9669524 |
| 10pp | cortex | 0.0035723 | 0.0067828 | 0.5266659 | 0.8289377 |
| 11l | cortex | 0.0111894 | 0.0067824 | 1.6497671 | 0.3428508 |
| 13l | cortex | 0.0103882 | 0.0067825 | 1.5316300 | 0.3794707 |
| OFC | cortex | 0.0113444 | 0.0067826 | 1.6725642 | 0.3414714 |
| 47s | cortex | -0.0013689 | 0.0067828 | -0.2018194 | 0.9133209 |
| LIPd | cortex | -0.0146524 | 0.0067821 | -2.1604554 | 0.2129606 |
| 6a | cortex | -0.0046994 | 0.0067827 | -0.6928469 | 0.7326642 |
| i6-8 | cortex | -0.0070213 | 0.0067827 | -1.0351917 | 0.6182092 |
| s6-8 | cortex | -0.0086581 | 0.0067826 | -1.2765260 | 0.4964930 |
| 43 | cortex | -0.0074222 | 0.0067826 | -1.0942907 | 0.5753709 |
| OP4 | cortex | 0.0043168 | 0.0067828 | 0.6364406 | 0.7544673 |
| OP1 | cortex | -0.0015381 | 0.0067828 | -0.2267703 | 0.9051420 |
| OP2-3 | cortex | -0.0105290 | 0.0067824 | -1.5523959 | 0.3794707 |
| 52 | cortex | -0.0103839 | 0.0067830 | -1.5308791 | 0.3794707 |
| RI | cortex | -0.0135249 | 0.0067822 | -1.9941751 | 0.2571494 |
| PFcm | cortex | -0.0203349 | 0.0067814 | -2.9986249 | 0.0634650 |
| PoI2 | cortex | 0.0019446 | 0.0067828 | 0.2866935 | 0.8829476 |
| TA2 | cortex | 0.0007071 | 0.0067828 | 0.1042461 | 0.9579825 |
| FOP4 | cortex | -0.0144965 | 0.0067821 | -2.1374677 | 0.2175284 |
| MI | cortex | 0.0126769 | 0.0067823 | 1.8691247 | 0.2726984 |
| Pir | cortex | -0.0139087 | 0.0067822 | -2.0507778 | 0.2431033 |
| AVI | cortex | 0.0037609 | 0.0067828 | 0.5544765 | 0.8144461 |
| AAIC | cortex | -0.0024006 | 0.0067828 | -0.3539308 | 0.8712596 |
| FOP1 | cortex | -0.0025410 | 0.0067828 | -0.3746246 | 0.8712596 |
| FOP3 | cortex | -0.0056662 | 0.0067831 | -0.8353324 | 0.6860176 |
| FOP2 | cortex | -0.0065657 | 0.0067827 | -0.9680166 | 0.6501731 |
| PFt | cortex | -0.0097912 | 0.0067825 | -1.4435925 | 0.4154974 |
| AIP | cortex | -0.0122334 | 0.0067823 | -1.8037233 | 0.2943295 |
| EC | cortex | 0.0121856 | 0.0067823 | 1.7966700 | 0.2943295 |
| PreS | cortex | 0.0012400 | 0.0067828 | 0.1828174 | 0.9188180 |
| H | cortex | 0.0112494 | 0.0067824 | 1.6586141 | 0.3428508 |
| ProS | cortex | -0.0131159 | 0.0067822 | -1.9338654 | 0.2726984 |
| PeEc | cortex | 0.0027639 | 0.0067828 | 0.4074823 | 0.8712596 |
| STGa | cortex | -0.0126748 | 0.0067823 | -1.8688190 | 0.2726984 |
| PBelt | cortex | -0.0079694 | 0.0067826 | -1.1749776 | 0.5343226 |
| A5 | cortex | -0.0070177 | 0.0067827 | -1.0346589 | 0.6182092 |
| PHA1 | cortex | 0.0032222 | 0.0067828 | 0.4750504 | 0.8468744 |
| PHA3 | cortex | -0.0012818 | 0.0067828 | -0.1889738 | 0.9188180 |
| STSda | cortex | -0.0033831 | 0.0067828 | -0.4987745 | 0.8402390 |

**Table ST6:** **linear regression results at each brain area with BMI as the dependent variable and cortical thickness or subcortical volume as an independent variable** *(continued)*

| Brain area | layer | $\beta$ | SE | t-value | $P_{FDR}$ |
|---|---|---|---|---|---|
| STSdp | cortex | 0.0000911 | 0.0067828 | 0.0134254 | 0.9957617 |
| STSvp | cortex | 0.0021693 | 0.0067828 | 0.3198179 | 0.8743274 |
| TGd | cortex | -0.0053294 | 0.0067827 | -0.7857381 | 0.7140984 |
| TE1a | cortex | 0.0051170 | 0.0067827 | 0.7544161 | 0.7140984 |
| TE1p | cortex | 0.0000342 | 0.0067828 | 0.0050428 | 0.9959765 |
| TE2a | cortex | 0.0075660 | 0.0067826 | 1.1154988 | 0.5623793 |
| TF | cortex | 0.0154650 | 0.0067820 | 2.2802949 | 0.1833266 |
| TE2p | cortex | 0.0031807 | 0.0067828 | 0.4689323 | 0.8468744 |
| PHT | cortex | 0.0051210 | 0.0067827 | 0.7550067 | 0.7140984 |
| PH | cortex | -0.0127840 | 0.0067823 | -1.8849104 | 0.2726984 |
| TPOJ1 | cortex | -0.0094653 | 0.0067825 | -1.3955461 | 0.4413890 |
| TPOJ2 | cortex | -0.0185936 | 0.0067816 | -2.7417607 | 0.0955915 |
| TPOJ3 | cortex | -0.0079845 | 0.0067826 | -1.1771969 | 0.5343226 |
| DVT | cortex | -0.0161369 | 0.0067819 | -2.3793984 | 0.1622178 |
| PGp | cortex | -0.0027552 | 0.0067828 | -0.4062013 | 0.8712596 |
| IP2 | cortex | -0.0114892 | 0.0067824 | -1.6939758 | 0.3414714 |
| IP1 | cortex | -0.0085231 | 0.0067826 | -1.2566215 | 0.5055245 |
| IP0 | cortex | -0.0047627 | 0.0067827 | -0.7021832 | 0.7326642 |
| PFop | cortex | -0.0026610 | 0.0067828 | -0.3923232 | 0.8712596 |
| PF | cortex | -0.0104788 | 0.0067824 | -1.5449946 | 0.3794707 |
| PFm | cortex | -0.0061107 | 0.0067827 | -0.9009310 | 0.6547404 |
| PGi | cortex | -0.0177909 | 0.0067817 | -2.6233539 | 0.1018327 |
| PGs | cortex | -0.0104427 | 0.0067824 | -1.5396581 | 0.3794707 |
| V6A | cortex | -0.0048869 | 0.0067827 | -0.7204846 | 0.7277287 |
| VMV1 | cortex | -0.0185871 | 0.0067816 | -2.7407879 | 0.0955915 |
| VMV3 | cortex | 0.0048682 | 0.0067827 | 0.7177313 | 0.7277287 |
| PHA2 | cortex | -0.0011028 | 0.0067828 | -0.1625820 | 0.9252772 |
| V4t | cortex | -0.0000813 | 0.0067828 | -0.0119862 | 0.9957617 |
| FST | cortex | -0.0020949 | 0.0067828 | -0.3088499 | 0.8743274 |
| V3CD | cortex | -0.0007069 | 0.0067828 | -0.1042155 | 0.9579825 |
| LO3 | cortex | -0.0024573 | 0.0067828 | -0.3622796 | 0.8712596 |
| VMV2 | cortex | 0.0003991 | 0.0067828 | 0.0588364 | 0.9695882 |
| 31pd | cortex | -0.0148484 | 0.0067821 | -2.1893561 | 0.2055674 |
| 31a | cortex | -0.0011554 | 0.0067828 | -0.1703478 | 0.9240346 |
| VVC | cortex | 0.0127968 | 0.0067823 | 1.8868038 | 0.2726984 |
| 25 | cortex | 0.0113263 | 0.0067826 | 1.6698946 | 0.3414714 |
| s32 | cortex | 0.0142732 | 0.0067821 | 2.1045260 | 0.2279081 |
| pOFC | cortex | 0.0031432 | 0.0067828 | 0.4634017 | 0.8468744 |
| PoI1 | cortex | -0.0039267 | 0.0067828 | -0.5789183 | 0.8031725 |
| Ig | cortex | -0.0065558 | 0.0067827 | -0.9665522 | 0.6501731 |
| FOP5 | cortex | -0.0044274 | 0.0067828 | -0.6527479 | 0.7508134 |
| p10p | cortex | -0.0055113 | 0.0067827 | -0.8125488 | 0.6953824 |
| p47r | cortex | -0.0178048 | 0.0067817 | -2.6253955 | 0.1018327 |
| TGv | cortex | 0.0023276 | 0.0067828 | 0.3431547 | 0.8712596 |
| MBelt | cortex | 0.0024815 | 0.0067828 | 0.3658529 | 0.8712596 |

**Table ST6:** **linear regression results at each brain area with BMI as the dependent variable and cortical thickness or subcortical volume as an independent variable** *(continued)*

| Brain area | layer | $\beta$ | SE | t-value | $P_{FDR}$ |
|---|---|---|---|---|---|
| LBelt | cortex | -0.0016323 | 0.0067828 | -0.2406532 | 0.9014136 |
| A4 | cortex | -0.0060323 | 0.0067827 | -0.8893734 | 0.6594612 |
| STSva | cortex | -0.0017739 | 0.0067828 | -0.2615279 | 0.8995130 |
| TE1m | cortex | 0.0110191 | 0.0067824 | 1.6246650 | 0.3544448 |
| PI | cortex | 0.0063200 | 0.0067827 | 0.9317801 | 0.6504152 |
| a32pr | cortex | -0.0014223 | 0.0067828 | -0.2096861 | 0.9119418 |
| p24 | cortex | 0.0165307 | 0.0067819 | 2.4374747 | 0.1456455 |

**Table ST7:** **linear regression results at each brain area with AT as the dependent variable and cortical thickness or subcortical volume as an independent variable**

| Brain area | layer | $\beta$ | SE | t-value | $P_{FDR}$ |
|---|---|---|---|---|---|
| Thalamus | subcortex | -0.0343446 | 0.0067791 | -5.0662711 | 0.0000361 |
| Caudate | subcortex | -0.0134896 | 0.0067838 | -1.9885112 | 0.2571494 |
| Putamen | subcortex | -0.0240733 | 0.0067820 | -3.5496031 | 0.0144598 |
| Pallidum | subcortex | -0.0427623 | 0.0067768 | -6.3101038 | 0.0000001 |
| Hippocampus | subcortex | -0.0275440 | 0.0067802 | -4.0623839 | 0.0022788 |
| Amygdala | subcortex | -0.0225451 | 0.0067811 | -3.3247045 | 0.0276320 |
| Accumbens | subcortex | -0.0338903 | 0.0067789 | -4.9993629 | 0.0000361 |
| V1 | cortex | -0.0026823 | 0.0067828 | -0.3954540 | 0.8712596 |
| MST | cortex | -0.0061870 | 0.0067827 | -0.9121820 | 0.6504152 |
| V6 | cortex | -0.0113643 | 0.0067824 | -1.6755612 | 0.3414714 |
| V2 | cortex | -0.0075707 | 0.0067826 | -1.1161897 | 0.5623793 |
| V3 | cortex | -0.0077400 | 0.0067826 | -1.1411486 | 0.5584055 |
| V4 | cortex | -0.0087003 | 0.0067826 | -1.2827432 | 0.4964930 |
| V8 | cortex | 0.0051557 | 0.0067827 | 0.7601246 | 0.7140984 |
| 4 | cortex | -0.0092737 | 0.0067825 | -1.3672875 | 0.4468037 |
| 3b | cortex | -0.0108675 | 0.0067846 | -1.6017949 | 0.3647018 |
| FEF | cortex | -0.0052475 | 0.0067827 | -0.7736608 | 0.7140984 |
| PEF | cortex | -0.0083379 | 0.0067826 | -1.2293058 | 0.5055245 |
| 55b | cortex | -0.0063611 | 0.0067827 | -0.9378386 | 0.6504152 |
| V3A | cortex | -0.0003910 | 0.0067828 | -0.0576433 | 0.9695882 |
| RSC | cortex | -0.0019627 | 0.0067832 | -0.2893516 | 0.8829476 |
| POS2 | cortex | -0.0052331 | 0.0067827 | -0.7715347 | 0.7140984 |
| V7 | cortex | -0.0068088 | 0.0067827 | -1.0038553 | 0.6275631 |
| IPS1 | cortex | -0.0021771 | 0.0067828 | -0.3209802 | 0.8743274 |
| FFC | cortex | 0.0021131 | 0.0067828 | 0.3115376 | 0.8743274 |
| V3B | cortex | -0.0020956 | 0.0067828 | -0.3089526 | 0.8743274 |
| LO1 | cortex | 0.0056191 | 0.0067827 | 0.8284472 | 0.6863850 |
| LO2 | cortex | 0.0045209 | 0.0067827 | 0.6665328 | 0.7436970 |
| PIT | cortex | -0.0102186 | 0.0067825 | -1.5066175 | 0.3915817 |
| MT | cortex | -0.0015185 | 0.0067828 | -0.2238754 | 0.9051420 |
| A1 | cortex | 0.0092637 | 0.0067825 | 1.3658161 | 0.4468037 |
| PSL | cortex | -0.0134039 | 0.0067822 | -1.9763341 | 0.2571494 |
| SFL | cortex | -0.0064590 | 0.0067827 | -0.9522753 | 0.6504152 |
| PCV | cortex | -0.0100389 | 0.0067825 | -1.4801164 | 0.3954582 |
| STV | cortex | -0.0154606 | 0.0067820 | -2.2796464 | 0.1833266 |
| 7Pm | cortex | 0.0061865 | 0.0067827 | 0.9120961 | 0.6504152 |
| 7m | cortex | 0.0089561 | 0.0067825 | 1.3204583 | 0.4717857 |
| POS1 | cortex | -0.0063129 | 0.0067827 | -0.9307364 | 0.6504152 |
| 23d | cortex | -0.0075756 | 0.0067841 | -1.1166775 | 0.5623793 |
| v23ab | cortex | 0.0006429 | 0.0067828 | 0.0947776 | 0.9604448 |
| d23ab | cortex | -0.0043473 | 0.0067831 | -0.6408993 | 0.7544673 |
| 31pv | cortex | -0.0068322 | 0.0067827 | -1.0073058 | 0.6275631 |
| 5m | cortex | -0.0025308 | 0.0067828 | -0.3731237 | 0.8712596 |
| 5mv | cortex | -0.0191878 | 0.0067816 | -2.8294042 | 0.0873442 |
| 23c | cortex | -0.0191864 | 0.0067816 | -2.8291941 | 0.0873442 |

**Table ST7:** **linear regression results at each brain area with AT as the dependent variable and cortical thickness or subcortical volume as an independent variable** *(continued)*

| Brain area | layer | $\beta$ | SE | t-value | $P_{FDR}$ |
|---|---|---|---|---|---|
| 5L | cortex | -0.0135293 | 0.0067822 | -1.9948183 | 0.2571494 |
| 24dd | cortex | -0.0177978 | 0.0067817 | -2.6243752 | 0.1018327 |
| 24dv | cortex | -0.0095565 | 0.0067825 | -1.4089887 | 0.4368452 |
| 7AL | cortex | -0.0114492 | 0.0067824 | -1.6880866 | 0.3414714 |
| SCEF | cortex | -0.0046428 | 0.0067827 | -0.6845000 | 0.7326642 |
| 6ma | cortex | -0.0160117 | 0.0067819 | -2.3609321 | 0.1624036 |
| 7Am | cortex | -0.0026092 | 0.0067828 | -0.3846798 | 0.8712596 |
| 7PL | cortex | -0.0092632 | 0.0067825 | -1.3657506 | 0.4468037 |
| 7PC | cortex | -0.0038723 | 0.0067828 | -0.5709017 | 0.8047689 |
| LIPv | cortex | -0.0084362 | 0.0067826 | -1.2438057 | 0.5055245 |
| VIP | cortex | -0.0057625 | 0.0067827 | -0.8495840 | 0.6820270 |
| MIP | cortex | -0.0083808 | 0.0067826 | -1.2356382 | 0.5055245 |
| 1 | cortex | -0.0119166 | 0.0067823 | -1.7570086 | 0.3140423 |
| 2 | cortex | -0.0139923 | 0.0067822 | -2.0631020 | 0.2431033 |
| 3a | cortex | -0.0083857 | 0.0067826 | -1.2363539 | 0.5055245 |
| 6d | cortex | -0.0082262 | 0.0067826 | -1.2128409 | 0.5135746 |
| 6mp | cortex | -0.0129269 | 0.0067823 | -1.9059887 | 0.2726984 |
| 6v | cortex | -0.0004454 | 0.0067828 | -0.0656723 | 0.9695882 |
| p24pr | cortex | -0.0032370 | 0.0067828 | -0.4772356 | 0.8468744 |
| 33pr | cortex | 0.0100207 | 0.0067825 | 1.4774352 | 0.3954582 |
| a24pr | cortex | 0.0057384 | 0.0067827 | 0.8460303 | 0.6820270 |
| p32pr | cortex | -0.0036090 | 0.0067830 | -0.5320661 | 0.8289377 |
| a24 | cortex | 0.0166490 | 0.0067820 | 2.4548827 | 0.1456455 |
| d32 | cortex | -0.0068137 | 0.0067827 | -1.0045732 | 0.6275631 |
| 8BM | cortex | -0.0034552 | 0.0067828 | -0.5094139 | 0.8393925 |
| p32 | cortex | -0.0016495 | 0.0067828 | -0.2431885 | 0.9014136 |
| 10r | cortex | 0.0033628 | 0.0067832 | 0.4957591 | 0.8402390 |
| 47m | cortex | -0.0008294 | 0.0067828 | -0.1222725 | 0.9536834 |
| 8Av | cortex | -0.0024311 | 0.0067828 | -0.3584145 | 0.8712596 |
| 8Ad | cortex | -0.0100225 | 0.0067825 | -1.4777031 | 0.3954582 |
| 9m | cortex | -0.0058499 | 0.0067827 | -0.8624668 | 0.6788628 |
| 8BL | cortex | -0.0062599 | 0.0067827 | -0.9229206 | 0.6504152 |
| 9p | cortex | -0.0171253 | 0.0067818 | -2.5251813 | 0.1272800 |
| 10d | cortex | -0.0048741 | 0.0067828 | -0.7186020 | 0.7277287 |
| 8C | cortex | -0.0104541 | 0.0067824 | -1.5413506 | 0.3794707 |
| 44 | cortex | -0.0126243 | 0.0067823 | -1.8613656 | 0.2726984 |
| 45 | cortex | -0.0128380 | 0.0067823 | -1.8928741 | 0.2726984 |
| 47l | cortex | 0.0023660 | 0.0067828 | 0.3488218 | 0.8712596 |
| a47r | cortex | -0.0046431 | 0.0067827 | -0.6845403 | 0.7326642 |
| 6r | cortex | -0.0217190 | 0.0067812 | -3.2028126 | 0.0364085 |
| IFJa | cortex | -0.0017187 | 0.0067828 | -0.2533946 | 0.9011662 |
| IFJp | cortex | 0.0025317 | 0.0067828 | 0.3732496 | 0.8712596 |
| IFSp | cortex | -0.0048479 | 0.0067827 | -0.7147441 | 0.7277287 |
| IFSa | cortex | -0.0124491 | 0.0067823 | -1.8355367 | 0.2823694 |
| p9-46v | cortex | -0.0090460 | 0.0067825 | -1.3337129 | 0.4670183 |

**Table ST7:** **linear regression results at each brain area with AT as the dependent variable and cortical thickness or subcortical volume as an independent variable** *(continued)*

| Brain area | layer | $\beta$ | SE | t-value | $P_{FDR}$ |
|---|---|---|---|---|---|
| 46 | cortex | -0.0150781 | 0.0067820 | -2.2232313 | 0.1960541 |
| a9-46v | cortex | -0.0063168 | 0.0067827 | -0.9313165 | 0.6504152 |
| 9-46d | cortex | -0.0179443 | 0.0067817 | -2.6459791 | 0.1018327 |
| 9a | cortex | -0.0153606 | 0.0067820 | -2.2649011 | 0.1833266 |
| 10v | cortex | 0.0026768 | 0.0067828 | 0.3946474 | 0.8712596 |
| a10p | cortex | 0.0005453 | 0.0067830 | 0.0803908 | 0.9669524 |
| 10pp | cortex | 0.0035723 | 0.0067828 | 0.5266659 | 0.8289377 |
| 11l | cortex | 0.0111894 | 0.0067824 | 1.6497671 | 0.3428508 |
| 13l | cortex | 0.0103882 | 0.0067825 | 1.5316300 | 0.3794707 |
| OFC | cortex | 0.0113444 | 0.0067826 | 1.6725642 | 0.3414714 |
| 47s | cortex | -0.0013689 | 0.0067828 | -0.2018194 | 0.9133209 |
| LIPd | cortex | -0.0146524 | 0.0067821 | -2.1604554 | 0.2129606 |
| 6a | cortex | -0.0046994 | 0.0067827 | -0.6928469 | 0.7326642 |
| i6-8 | cortex | -0.0070213 | 0.0067827 | -1.0351917 | 0.6182092 |
| s6-8 | cortex | -0.0086581 | 0.0067826 | -1.2765260 | 0.4964930 |
| 43 | cortex | -0.0074222 | 0.0067826 | -1.0942907 | 0.5753709 |
| OP4 | cortex | 0.0043168 | 0.0067828 | 0.6364406 | 0.7544673 |
| OP1 | cortex | -0.0015381 | 0.0067828 | -0.2267703 | 0.9051420 |
| OP2-3 | cortex | -0.0105290 | 0.0067824 | -1.5523959 | 0.3794707 |
| 52 | cortex | -0.0103839 | 0.0067830 | -1.5308791 | 0.3794707 |
| RI | cortex | -0.0135249 | 0.0067822 | -1.9941751 | 0.2571494 |
| PFcm | cortex | -0.0203349 | 0.0067814 | -2.9986249 | 0.0634650 |
| PoI2 | cortex | 0.0019446 | 0.0067828 | 0.2866935 | 0.8829476 |
| TA2 | cortex | 0.0007071 | 0.0067828 | 0.1042461 | 0.9579825 |
| FOP4 | cortex | -0.0144965 | 0.0067821 | -2.1374677 | 0.2175284 |
| MI | cortex | 0.0126769 | 0.0067823 | 1.8691247 | 0.2726984 |
| Pir | cortex | -0.0139087 | 0.0067822 | -2.0507778 | 0.2431033 |
| AVI | cortex | 0.0037609 | 0.0067828 | 0.5544765 | 0.8144461 |
| AAIC | cortex | -0.0024006 | 0.0067828 | -0.3539308 | 0.8712596 |
| FOP1 | cortex | -0.0025410 | 0.0067828 | -0.3746246 | 0.8712596 |
| FOP3 | cortex | -0.0056662 | 0.0067831 | -0.8353324 | 0.6860176 |
| FOP2 | cortex | -0.0065657 | 0.0067827 | -0.9680166 | 0.6501731 |
| PFt | cortex | -0.0097912 | 0.0067825 | -1.4435925 | 0.4154974 |
| AIP | cortex | -0.0122334 | 0.0067823 | -1.8037233 | 0.2943295 |
| EC | cortex | 0.0121856 | 0.0067823 | 1.7966700 | 0.2943295 |
| PreS | cortex | 0.0012400 | 0.0067828 | 0.1828174 | 0.9188180 |
| H | cortex | 0.0112494 | 0.0067824 | 1.6586141 | 0.3428508 |
| ProS | cortex | -0.0131159 | 0.0067822 | -1.9338654 | 0.2726984 |
| PeEc | cortex | 0.0027639 | 0.0067828 | 0.4074823 | 0.8712596 |
| STGa | cortex | -0.0126748 | 0.0067823 | -1.8688190 | 0.2726984 |
| PBelt | cortex | -0.0079694 | 0.0067826 | -1.1749776 | 0.5343226 |
| A5 | cortex | -0.0070177 | 0.0067827 | -1.0346589 | 0.6182092 |
| PHA1 | cortex | 0.0032222 | 0.0067828 | 0.4750504 | 0.8468744 |
| PHA3 | cortex | -0.0012818 | 0.0067828 | -0.1889738 | 0.9188180 |
| STSda | cortex | -0.0033831 | 0.0067828 | -0.4987745 | 0.8402390 |

**Table ST7:** **linear regression results at each brain area with AT as the dependent variable and cortical thickness or subcortical volume as an independent variable** *(continued)*

| Brain area | layer | $\beta$ | SE | t-value | $P_{FDR}$ |
|---|---|---|---|---|---|
| STSdp | cortex | 0.0000911 | 0.0067828 | 0.0134254 | 0.9957617 |
| STSvp | cortex | 0.0021693 | 0.0067828 | 0.3198179 | 0.8743274 |
| TGd | cortex | -0.0053294 | 0.0067827 | -0.7857381 | 0.7140984 |
| TE1a | cortex | 0.0051170 | 0.0067827 | 0.7544161 | 0.7140984 |
| TE1p | cortex | 0.0000342 | 0.0067828 | 0.0050428 | 0.9959765 |
| TE2a | cortex | 0.0075660 | 0.0067826 | 1.1154988 | 0.5623793 |
| TF | cortex | 0.0154650 | 0.0067820 | 2.2802949 | 0.1833266 |
| TE2p | cortex | 0.0031807 | 0.0067828 | 0.4689323 | 0.8468744 |
| PHT | cortex | 0.0051210 | 0.0067827 | 0.7550067 | 0.7140984 |
| PH | cortex | -0.0127840 | 0.0067823 | -1.8849104 | 0.2726984 |
| TPOJ1 | cortex | -0.0094653 | 0.0067825 | -1.3955461 | 0.4413890 |
| TPOJ2 | cortex | -0.0185936 | 0.0067816 | -2.7417607 | 0.0955915 |
| TPOJ3 | cortex | -0.0079845 | 0.0067826 | -1.1771969 | 0.5343226 |
| DVT | cortex | -0.0161369 | 0.0067819 | -2.3793984 | 0.1622178 |
| PGp | cortex | -0.0027552 | 0.0067828 | -0.4062013 | 0.8712596 |
| IP2 | cortex | -0.0114892 | 0.0067824 | -1.6939758 | 0.3414714 |
| IP1 | cortex | -0.0085231 | 0.0067826 | -1.2566215 | 0.5055245 |
| IP0 | cortex | -0.0047627 | 0.0067827 | -0.7021832 | 0.7326642 |
| PFop | cortex | -0.0026610 | 0.0067828 | -0.3923232 | 0.8712596 |
| PF | cortex | -0.0104788 | 0.0067824 | -1.5449946 | 0.3794707 |
| PFm | cortex | -0.0061107 | 0.0067827 | -0.9009310 | 0.6547404 |
| PGi | cortex | -0.0177909 | 0.0067817 | -2.6233539 | 0.1018327 |
| PGs | cortex | -0.0104427 | 0.0067824 | -1.5396581 | 0.3794707 |
| V6A | cortex | -0.0048869 | 0.0067827 | -0.7204846 | 0.7277287 |
| VMV1 | cortex | -0.0185871 | 0.0067816 | -2.7407879 | 0.0955915 |
| VMV3 | cortex | 0.0048682 | 0.0067827 | 0.7177313 | 0.7277287 |
| PHA2 | cortex | -0.0011028 | 0.0067828 | -0.1625820 | 0.9252772 |
| V4t | cortex | -0.0000813 | 0.0067828 | -0.0119862 | 0.9957617 |
| FST | cortex | -0.0020949 | 0.0067828 | -0.3088499 | 0.8743274 |
| V3CD | cortex | -0.0007069 | 0.0067828 | -0.1042155 | 0.9579825 |
| LO3 | cortex | -0.0024573 | 0.0067828 | -0.3622796 | 0.8712596 |
| VMV2 | cortex | 0.0003991 | 0.0067828 | 0.0588364 | 0.9695882 |
| 31pd | cortex | -0.0148484 | 0.0067821 | -2.1893561 | 0.2055674 |
| 31a | cortex | -0.0011554 | 0.0067828 | -0.1703478 | 0.9240346 |
| VVC | cortex | 0.0127968 | 0.0067823 | 1.8868038 | 0.2726984 |
| 25 | cortex | 0.0113263 | 0.0067826 | 1.6698946 | 0.3414714 |
| s32 | cortex | 0.0142732 | 0.0067821 | 2.1045260 | 0.2279081 |
| pOFC | cortex | 0.0031432 | 0.0067828 | 0.4634017 | 0.8468744 |
| PoI1 | cortex | -0.0039267 | 0.0067828 | -0.5789183 | 0.8031725 |
| Ig | cortex | -0.0065558 | 0.0067827 | -0.9665522 | 0.6501731 |
| FOP5 | cortex | -0.0044274 | 0.0067828 | -0.6527479 | 0.7508134 |
| p10p | cortex | -0.0055113 | 0.0067827 | -0.8125488 | 0.6953824 |
| p47r | cortex | -0.0178048 | 0.0067817 | -2.6253955 | 0.1018327 |
| TGv | cortex | 0.0023276 | 0.0067828 | 0.3431547 | 0.8712596 |
| MBelt | cortex | 0.0024815 | 0.0067828 | 0.3658529 | 0.8712596 |

**Table ST7:** **linear regression results at each brain area with AT as the dependent variable and cortical thickness or subcortical volume as an independent variable** *(continued)*

| **Brain area** | **layer** | $\beta$ | **SE** | **t-value** | $P_{FDR}$ |
|---|---|---|---|---|---|
| LBelt | cortex | -0.0016323 | 0.0067828 | -0.2406532 | 0.9014136 |
| A4 | cortex | -0.0060323 | 0.0067827 | -0.8893734 | 0.6594612 |
| STSva | cortex | -0.0017739 | 0.0067828 | -0.2615279 | 0.8995130 |
| TE1m | cortex | 0.0110191 | 0.0067824 | 1.6246650 | 0.3544448 |
| PI | cortex | 0.0063200 | 0.0067827 | 0.9317801 | 0.6504152 |
| a32pr | cortex | -0.0014223 | 0.0067828 | -0.2096861 | 0.9119418 |
| p24 | cortex | 0.0165307 | 0.0067819 | 2.4374747 | 0.1456455 |
